# Fusion oncoproteins and cooperating mutations define disease phenotypes in *NUP98*-rearranged leukemia

**DOI:** 10.1101/2025.01.21.25320683

**Authors:** Masayuki Umeda, Ryan Hiltenbrand, Nicole L. Michmerhuizen, Juan M. Barajas, Melvin E. Thomas, Bright Arthur, Michael P Walsh, Guangchun Song, Jing Ma, Tamara Westover, Amit Kumar, Petri Pölönen, Cristina Mecucci, Danika Di Giacomo, Franco Locatelli, Riccardo Masetti, Salvatore N. Bertuccio, Martina Pigazzi, Shondra M. Pruett-Miller, Stanley Pounds, Jeffrey Rubnitz, Hiroto Inaba, Kyriakos P. Papadopoulos, Michael J. Wick, Ilaria Iacobucci, Charles G. Mullighan, Jeffery M. Klco

## Abstract

Leukemias with *NUP98* rearrangements exhibit heterogeneous phenotypes correlated to fusion partners, whereas the mechanism responsible for this heterogeneity is poorly understood. Through genome-wide mutational and transcriptional analyses of 177 *NUP98*-rearranged leukemias, we show that cooperating alterations are associated with differentiation status even among leukemias sharing the same *NUP98* fusions, such as *NUP98::KDM5A* acute megakaryocytic leukemia with *RB1* loss or T-cell acute lymphoblastic leukemia with *NOTCH1* mutations. CUT&RUN profiling reveals that NUP98 fusion oncoproteins directly regulate differentiation-related genes, with binding patterns also influenced by differentiation stage. Using *in vitro* models, we show *RB1* loss cooperates with NUP98::KDM5A by blocking terminal differentiation toward platelets and expanding megakaryocyte-like cells, whereas *WT1* frameshifts skew differentiation toward dormant lympho-myeloid primed progenitor cells and cycling granulocyte-monocyte progenitor cells. NUP98::KDM5A models with *RB1* or *WT1* alterations have different sensitivities to menin inhibition, suggesting cellular differentiation stage-specific resistant mechanism against menin inhibitors with clinical implications for *NUP98*-rearranged leukemia.

## Introduction

The prognosis of children with acute myeloid leukemia (AML) remains unsatisfactory with high rates of disease relapse^1–3^. This is partly attributed to pediatric AML subtypes associated with poor treatment response, such as *KMT2A* rearrangement (*KMT2A*r)^4,5^, *UBTF* tandem duplications (*UBTF*-TD)^6^, *CBFA2T3*::*GLIS2*^7,8^, and *NUP98* rearrangement (*NUP98*r)^5,9,10^. Understanding the genetic background and disease mechanisms of these subtypes is needed to develop efficient treatment strategies based on biology and molecular mechanisms.

*NUP98*r with various fusion partners are known to be associated with specific hematologic malignancies^11^. For example, *NUP98::NSD1* is predominantly found in adolescents with AML, whereas *NUP98::KDM5A* is often found in acute megakaryocytic leukemia (AMKL) in young children^5,8,9^. Many other *NUP98*r with a variety of fusion partners have been reported in other hematological malignancies, such as T-cell acute lymphoblastic leukemia (T-ALL)^12^, myelodysplastic neoplasms (MDS)^13^ or mixed phenotype acute leukemia (MPAL)^14^. Recent genomic studies focusing on *NUP98*r AML confirmed previously known associations of *NUP98::NSD1* with *FLT3* internal tandem duplications (ITD) and *WT1* mutations or *NUP98::KDM5A* with chromosome 13 loss involving the *RB1* locus^8,10,15^. However, more comprehensive and genome-wide studies on various *NUP98*r leukemias are needed to better understand the association of genomic features, including cooperating mutations, with disease phenotypes.

From a mechanistic standpoint, recent studies revealed that NUP98 fusion oncoproteins (FOs) directly bind to gene loci expressed in *NUP98*r AML, such as *MEIS1*, *HOXA/B* clusters, and *CDK6*, with some similarities to AMLs with *NPM1* mutations or *KMT2A* rearrangements^16–18^. This binding is partly driven by intrinsically disordered regions of the N-terminal NUP98, which mediate transcriptional condensates^19,20^, and DNA/chromatin binding domains of the C-terminal fusion partners, leading to target gene activation. This knowledge has mostly been derived from mouse or non-hematopoietic human models due to a lack of appropriate human *NUP98*r leukemia models. An understanding of molecular mechanisms of leukemogenesis by NUP98 FOs in human hematopoietic cells, including the association of genomic binding with histone modification and transcriptional profiles, is necessary for developing mechanism-based novel therapeutics for *NUP98*r leukemias.

In this study, we performed genome-wide genetic characterization of 177 pediatric and young adult *NUP98*r hematologic malignancies including AML and T-ALL, using RNA sequencing (RNAseq) and whole genome/exome sequencing (WGS/WES), to reveal phenotype-specific mutational profiles. These data were corroborated by single cell transcriptomics and epigenetic profiling by CUT&RUN^21^, revealing NUP98-FO binding profiles associated with fusion partners and disease phenotypes of patient samples. Functional characterization of recurrent gene alterations using cord blood CD34+ cell models showed the NUP98 fusion- and phenotype-dependent impact on cell growth and transcriptional profiles associated with drug responses, which can be a basis for overcoming these refractory diseases.

## Results

### Transcriptional heterogeneity of pediatric NUP98r leukemia

We first established a pediatric and young adult *NUP98*r leukemia cohort with 185 samples from 177 patients (median age: 8.6, range: 0.4-28) by collecting new cases and integrating data across published cohorts^3,6,8,10,13,22-30^ (Fig.S1A, Supplementary Table 1), including 8 patients with data at multiple time points (Fig.1A). AML was the dominant disease (n=162), followed by T-ALL (n=18), therapy-related myeloid neoplasms (t-MN, n=4), and MDS (n=1), reflecting the heterogeneity of *NUP98*r hematologic neoplasms. We obtained RNAseq data for all samples (n=185) and DNA data for 91 samples (WGS:43, WES:16, both:32, Supplementary Tables 2-9). Fusion genes were identified by CICERO^31^ and manual inspection of sequencing data, confirming *NUP98* fusion transcripts in all cases.

**Figure 1.**
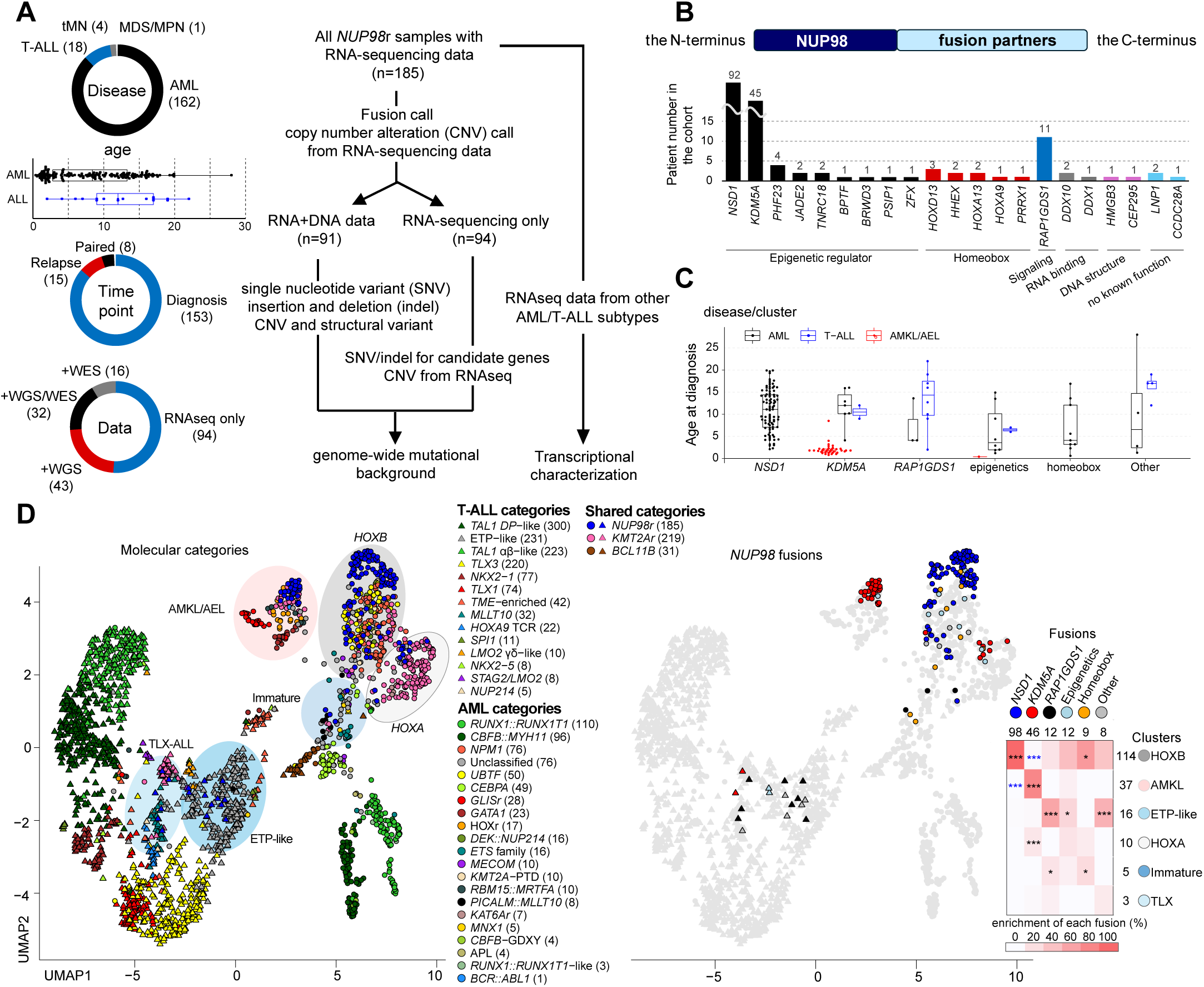
Heterogeneity of pediatric NUP98r leukemia. **A.** Details of *NUP98*-rearranged (*NUP98*r) leukemia samples (n=185, **left**) and analytical pipelines (**right**). **B.** Numbers and functional annotations of fusion partners in the study cohort. Colors indicate protein functional groups. **C**. Age distribution related to fusion partners and disease types. Colors indicate disease types. **D**. UMAP (Uniform Manifold Approximation and Projection) plots of transcriptional cohort (n=2,321) colored according to leukemia subtypes (**left**) and *NUP98* fusion partners (**mid**) and enrichment of fusion partners in transcriptional clusters (**right**). The shapes of dots indicate disease types (circles-acute myeloid leukemia: AML, triangles-acute lymphoblastic leukemia: ALL), and colors in the heatmap indicate enrichment of fusions in each cluster, asterisks indicate statistically significant adjusted *P*-values from two-sided Fisher’s exact tests and the Benjamini-Hochberg adjustment (*<0.05, **<0.01, ***<0.001, black: enriched, blue: exclusive). In **A** and **C**, lines of the box plots represent 25% quantile, median, and 75% quantile and the upper whisker represents the higher value of maxima or 1.5 x interquartile range (IQR), and the lower whisker represents the lower value of minima or 1.5 x IQR. Abbreviations. tMN: therapy-related myeloid neoplasm, MDS/MPN: myelodysplastic syndrome/ myeloproliferative neoplasms, AEL/AMKL: acute erythroid leukemia/ acute megakaryocytic leukemia, ETP: Early T-cell precursor ALL. Abbreviations in leukemia subtypes are found in Table S10.

Among fusion partners, *NSD1* was the most frequent (n=92, 51.7%), followed by *KDM5A* (n=45, 25.3%, Fig.1B, Fig.S1B, Supplementary Table 8). These fusions were enriched in specific age groups (Fig.1C). *RAP1GDS1* (n=11, 6.2%) was the third most common partner, with nine *NUP98::RAP1GDS1* were found in T-ALL cases. Most of the other partners fall into functional categories of epigenetic regulators (n=12, 6.7%) or homeobox proteins (n=9, 5.1%).

The majority of the partners with epigenetic regulatory functions, including NSD1 and KDM5A, had plant homeodomain (PHD) fingers in the retained C-terminus, suggesting structural redundancy among fusion partners (Fig.S1C). Functions of the remaining 6 partners (8 patients) were broad, ranging from RNA-binding (DDX1/10), maintenance of DNA structure (HMGB3 and CEP295), or without known function (LNP1 and CCDC28A), but all these partners had domains with multiple α-helices with linker sequences similar to homeobox domains or PHD. *NUP98* breakpoints were heterogeneous, ranging from exon 7-15, although fusion partners were strongly associated with specific exons (*RAP1GDS1-*exon 11, *NSD1-*exon 12, and *KDM5A-*exon 13, Fig.S1B).

Appreciating the known heterogeneity of the disease phenotypes and transcriptional profiles of *NUP98*r leukemia, we performed transcriptional analyses with other subtypes of AML^5^ (n=816) and T-ALL^12^ (n=1,320) as a background cohort (Fig.S2A, Supplementary Table 10). UMAP (Uniform Manifold Approximation and Projection) and unsupervised clustering revealed 10 clusters driven by driver alterations and differentiation status. *NUP98*r malignancies were distributed to 6 of these clusters (Fig.1D, Fig.S2B-D). The majority of *NUP98::NSD1* samples were clustered with AML with *NPM1* mutations or *UBTF*-TD showing high *HOXA/B* gene expression (the *HOXB* cluster). By contrast, samples with *NUP98::KDM5A* were predominantly found in a cluster with other AMKL subtypes (the AMKL cluster), although rare cases clustered with subtypes with *KMT2A*-rearranged and *KAT6A*-rearranged AML with *HOXA* expression (the *HOXA* cluster) or with mature ALL (the *TLX* cluster). *NUP98*::*RAP1GDS1* samples were mainly clustered with other early T-cell precursor-like ALL (the ETP-like cluster), while they were also found in the *HOXB*, *TLX*, or immature AML clusters. Although *HOXA-B* gene expression is recognized as a hallmark of *NUP98*r leukemia, we observed that *NUP98::NSD1* samples lack posterior *HOXB* (*HOXB6-9*) expression and immature/T-ALL cases lack *HOXB* or both *HOXA-B* expression (Fig.S2E). These data show that *NUP98*r leukemias are heterogeneous and even the same fusion partners can contribute to variable phenotypes of these hematopoietic malignancies.

### Mutational background of NUP98r leukemia associated with disease phenotypes

To assess genetic factors contributing to the transcriptional heterogeneity of *NUP98*r, we investigated the genomic landscape using DNA data and an RNAseq-based pipeline^5,6^ (see Methods), including single nucleotide variants (SNV), insertions and deletions (indel), ITD, copy number variations (CNV), and fusion genes other than *NUP98* rearrangements (Supplementary Tables 5-9). Among 53 recurrently altered genes, the most frequent alteration was *FLT3*-ITD (n=70, 37.8%), followed by *WT1* (n=44, 23.8%) and *NRAS* mutations (n=30, 16.2%, Fig.2A-C). Chromosome 13 deletions involving *RB1*^8,10^ were found predominantly in *NUP98::KDM5A* cases (n=23, 14.1%), with deleted regions extending in both 3’ and 5’ directions of *RB1*, often affecting the tumor suppressor *BRCA2* (Fig.S3A). We also identified *RB1* loss in cases with only RNAseq by assessing for allelic imbalance of single nucleotide polymorphisms (SNPs) within and around the *RB1* locus; however, *RB1* loss could still be underestimated as AMKL cases without detectable CNV also showed low *RB1* RNA expression (Fig.S3B-C). *RB1* loss was associated with high expression of stem cell markers (*GATA2, CD96,* and *BMI1*) among *NUP98::KDM5A* AMKL samples, and gene set enrichment analysis (GSEA) showed enrichment of genes involving AMKL phenotypes (Fig.S3D-F). GSEA also showed changes in genes related to cell growth and mitochondrial function, which are consistent with the known function of RB1/E2F^32,33^ (Fig.S3F). Additionally, genome-wide profiling revealed *JAK2* (n=6, 3.2%) mutations enriched in *NUP98::KDM5A* and the AMKL cluster, while *GATA1* (n=3, 1.6%) and *MPL* (n=3) mutations were associated only with the AMKL cluster (Fig.2C), all of which were frequently found in non-Down syndrome AMKL^8^.

**Figure 2.**
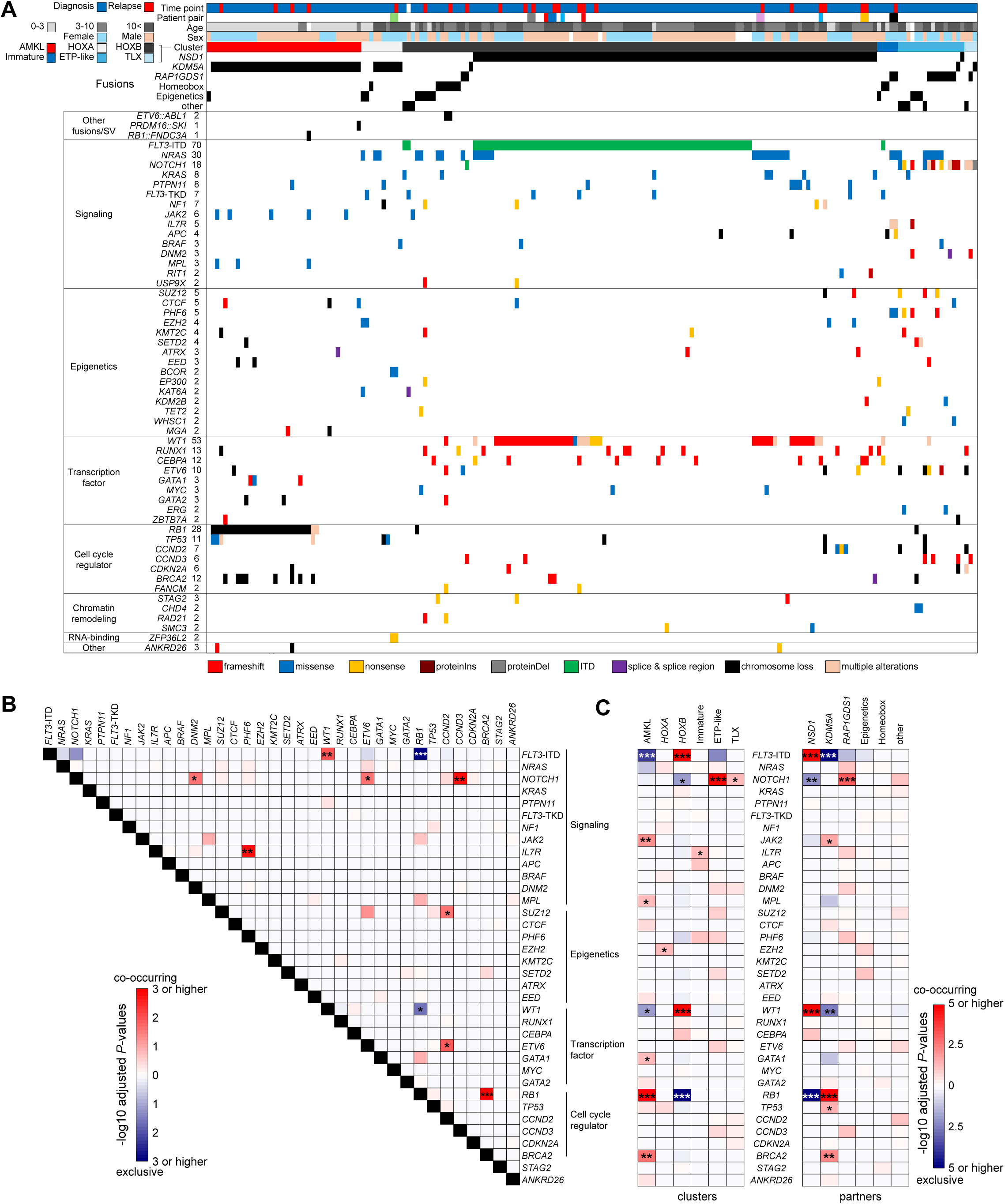
Mutational background of NUP98r leukemia associated with disease phenotypes. **A**. Genetic profiles of *NUP98*r samples in the cohort. Colors indicate patient annotations (**top**) and types of gene alterations (**bottom**). **B**. Co-occurrence and mutual exclusivity among recurrent alterations (n≥3). **C.** Enrichment of somatic alterations in transcriptional clusters (**left**) and fusion partners (**right**). In **B** and **C**, colors indicate adjusted *P*-values by two-sided Fisher’s exact tests and the Benjamini-Hochberg adjustment (red: co-occurring, blue: mutually exclusive), and asterisks indicate statistically significant values (adjusted *P*-values *<0.05, **<0.01, ***<0.001). Annotations of genes in mutational heatmaps depend on known general functions.

*FLT3*-ITD and *WT1* were highly co-occurring and enriched in *NUP98::NSD1* AMLs (Fig.2A-C). Variant allele frequencies (VAFs) of *WT1* and *FLT3*-ITD were generally high both at diagnosis and relapse (Fig.S4A-C). *FLT3*-ITD and *RAS*-related mutations were less likely to co-occur, especially among *NUP98*::*NSD1* samples, and VAFs of *RAS* mutations were more variable (Fig.2B, Fig.S4A-B), indicating that *RAS* mutations could exist in independent clones or subclonal to *FLT3*-ITD^34^. Differentially expressed gene (DEG) analysis showed high expression of immunity-related genes, such as *MRC1* (CD206) or *IL2RA* (CD25), in *FLT3*-ITD+ *NUP98*::*NSD1* AML, consistent with previous finding in *FLT3*-ITD+ AML, where these markers are associated with inferior outcomes^35,36^. GSEA confirmed known enrichment of inflammatory-related genes^37^ (Fig.S4D-E). Samples with *WT1* mutations had significantly high expression of genes such as *DNTT* (TdT: Terminal deoxynucleotidyl transferase), which is a known marker for lymphoid-myeloid primed progenitors (LMPP)^38,39^, and gene sets related to mitochondrial function and translation by ribosomes. These data suggest that secondary alterations in *NUP98::NSD1* AMLs can influence expression profiles and differentiation status.

Clinically, all 12 *NUP98::RAP1GDS1* leukemias were either T-ALL and immature AML (FAB M0-1). Those with *NOTCH1* mutations (n=7, 58.3%) were predominantly found in the *TLX* and ETP-like clusters (6/7, 85.7%) and associated with T cell-related expression profiles, whereas *NRAS* mutations were mainly found in the ETP and immature AML clusters (Fig.S4F-H). Mutational profiling of samples in T-ALL/immature AML clusters showed *NOTCH1* mutations enriched in the ALL clusters, including those in *NUP98::KDM5A* ALL, whereas *IL7R* mutations were enriched in immature AML clusters (Fig.2C). Thus, among immature *NUP98*r leukemias, *NOTCH1* mutations likely contribute to the establishment of T-ALL phenotypes^40^. These data show the strong association among fusion partners, cooperating mutations, and disease phenotypes of *NUP98*r malignancies.

### Variety of cellular hierarchies in NUP98r leukemia associated with fusions and cooperating alterations

Recent research showed associations between differentiation status or cellular hierarchies with outcomes or drug responses in AML^41–43^. To investigate the differentiation status of *NUP98*r leukemia, we performed single cell RNAseq (scRNAseq) on 9 representative samples from the cohort and obtained publicly available data from one case with diagnosis-relapse paired (D-R) samples of *NUP98::NSD1* AML (PAXLWH)^43^ (Fig.3A, Fig.S5A, Supplementary Table 11). UMAP analysis showed distinct clusters for each patient sample indicating unique expression profiles of each tumor, whereas T-cell or B-cell clusters consisted of cells from multiple patients reflecting normal lymphocytes, as reported previously^43,44^ (Fig.3B-C, Fig.S5B). To infer the differentiation stages compared to normal hematopoiesis, we projected scRNAseq data onto reference normal bone marrow and thymocyte scRNAseq data^44^, which revealed unique distributions of tumor cells in each sample (Fig.3D-F, Fig.S5C). Three *NUP98::NSD1* samples (SJAML015363_D5, SJAML016582_D1, and SJAML061252_R1) showed various cellular hierarchies ranging from LMPP-like to monocyte-like cells (Fig.3E-F). SJAML001441 D-R pair (*FLT3*-ITD+/*WT1*+) had enriched monocyte and granulocyte-macrophage progenitor (GMP)-like cells at diagnosis with increasing LMPP-like cells at relapse with an additional *WT1* mutation. PAXLWH D-R pair (*FLT3*-ITD+) showed homogenous LMPP-like cells at diagnosis but increased GMP or monocytic populations at relapse with an acquired *RUNX1* mutation, suggesting different patterns of relapse of these *NUP98::NSD1* cases. DEG analyses between cells from *NUP98:NSD1* patients and normal counterparts showed aberrantly high expression of *HOXA/B* genes along with low expression of marker genes (*e.g.*, *S100A12* in monocyte *and ELANE* in GMP) in leukemic samples (Fig.S5D), confirming aberrant leukemia expression profiles. Hierarchies of bulk RNA samples inferred by deconvolution using CIBERSORT^45^ revealed heterogeneous hierarchies among the entire cohort and variable patterns among both D-R pairs and somatic mutations (Fig.3G, Fig.S5E). Relapse samples commonly showed low monocyte and high LMPP signatures, indicating expansion of stem-like populations at relapse in a subset of *NUP98::NSD1* AML as reported in other subtypes^43^.

**Figure 3.**
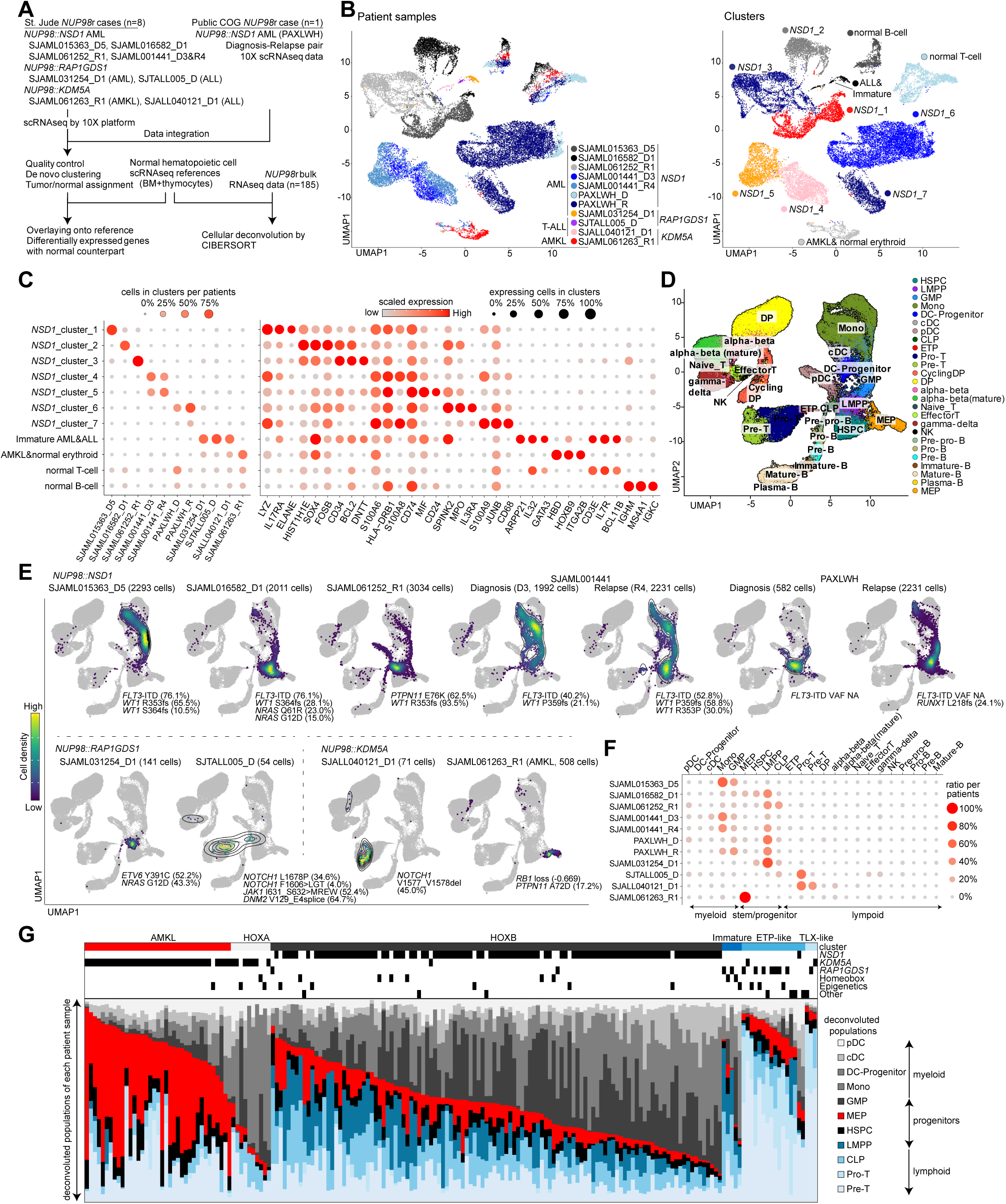
Variety of cellular hierarchies in NUP98r leukemia. **A**. Strategies for single cell RNAseq (scRNAseq) and deconvolution of bulk RNAseq data. **B**. UMAP plots of patient samples colored by sample source (**left**) and transcriptional clusters (**right**). **C**. Enrichment of cells in each cluster indicated by colors and sizes (**left**) and marker gene expression indicated by colors (averaged expression) and size (ratio of expressing cells: count>0). **D**. UMAP plot of reference bone marrow and thymocyte scRNAseq data, colored according to cell labels from the original reference data^44^. **E**. Distribution of patient sample scRNAseq on the reference data inferred by MapQuery function in Seurat package. Cells in normal hematopoietic cell clusters were excluded. Cells are colored according to the cell density on UMAP. Cooperating mutations found in bulk samples are also shown. **F**. Enrichment of cells with each cell label inferred by Seurat, indicated by colors and sizes. **G**. Cellular component of bulk RNA samples (n=185) inferred by CIBERSORT using signature matrix derived from reference scRNAseq data. Bars are colored by cell populations in each sample. Abbreviations. HSPC: hematopoietic stem and progenitor cell, LMPP: Lympho-myeloid primed progenitor, GMP: Granulocyte-monocyte progenitor, cDC: classic dendritic cell, pDC: plasmacytoid dendritic cell, CLP: common lymphoid progenitor, DP: CD4-CD8 double-positive T-cell, NK: natural killer T-cell, MEP: Megakaryocyte–erythroid progenitor

A *NUP98::KDM5A* AMKL case with *RB1* loss showed a cluster corresponding to megakaryocyte/erythrocyte progenitors (MEP) in the normal reference (Fig.3B,E-F). DEG analysis of the AMKL cluster with normal MEP revealed aberrant expressions of *HOXB* genes and immune response-related *IFI27* in AMKL cells, which were co-expressed at the single cell level (Fig.S6A-B). *NOTCH1^+^ NUP98::KDM5A* ALL cells were enriched at Pro- and Pre-T cell stages with aberrant expression of hemoglobin or *HOXB* genes (Fig.S6A).

*NOTCH1^+^ NUP98::RAP1GDS1* cells distributed broadly at the LMPP-ETP-Pro-T stages whereas *NUP98::RAP1GDS1* AML cells were enriched in the LMPP cluster (Fig.3E-F, Fig.S6C). Deconvoluted bulk *NUP98::RAP1GDS1* samples in immature clusters showed LMPP-CLP (common lymphoid progenitor) signatures, while those in the ALL clusters showed high Pro-Pre-T cell signatures, suggesting that the differentiation block of *NUP98::RAP1GDS1* occurs at various stages during T-cell development from hematopoietic stem cells (HSC), possibly affected by cooperating mutations^40^ (Fig.S6C-E). Two *NUP98::KDM5A* ALL cases showed higher Pre-T signals than *NUP98::RAP1GDS1* cases (Fig.S6F). These observations indicate that NUP98::KDM5A could contribute to the development of more mature ALL than NUP98::RAP1GDS1, despite both cooperating with *NOTCH1* mutations.

### Cord blood CD34 models recapitulate phenotypes of NUP98r leukemia

There are limited human *NUP98*r leukemia models that recapitulate the disease phenotypes^40,46–49^. To fill this gap, we tested the transforming potential of various NUP98 FOs by transducing cord blood CD34+ cells (cbCD34) with lentiviral particles encoding major NUP98-FOs (pediatrics-NUP98::NSD1 and NUP98:KDM5A, adult-NUP98::HOXA9). Expression of these NUP98-FOs increased clonogenic potential in methylcellulose and enhanced cell growth in liquid culture compared with an empty vector control (Fig.4A-C), despite a gradual decrease in CD34 expression. Flow cytometry analyses showed low CD11b expression in all conditions compared with the control, whereas only NUP98::KDM5A showed high CD41a expression, indicating preferential differentiation toward megakaryocytes (Fig.4D, Fig.S7A-B). We performed RNAseq from liquid cultures at multiple time points and observed dynamic changes of transcriptional status unique to each FO (Fig.4E-G, Fig.S7C-E). Consistent with patient samples, NUP98-FO-transduced conditions showed high *TAL1* and *MYCN* expression in addition to upregulation of *HOXA*/*B* genes, whereas control conditions showed reduced *TAL1* and increased *ITGAM* (CD11b) expression, suggesting myeloid differentiation (Fig.4F). We identified 320 common genes with high expression in conditions with NUP98 FOs, which enriched homeobox genes (Fig.4G). Notably, we also identified uniquely high genes with each FO related to specific functions, such as *GFI1B or GATA1* related to erythromegakaryocytic differentiation in cells with NUP98::KDM5A, consistent with surface markers. Comparison with the patient cohort showed clustering of cbCD34 models with patient samples with similar fusions/phenotypes (Fig.S7E). Two cell lines with *NUP98::KDM5A*, CHRF-288-11^46^ (megakaryocytic) and ST1653^50^ (myelomonocytic, patient-derived xenograft: PDX) also showed unique patterns of expression (e.g., *ITGA2B* and *CD34* in CHRF-288-11, and *ITGAM* in ST1653), confirming the heterogeneity of leukemia with *NUP98::KDM5A* (Fig.4E-F, Fig.S7E).

**Figure 4.**
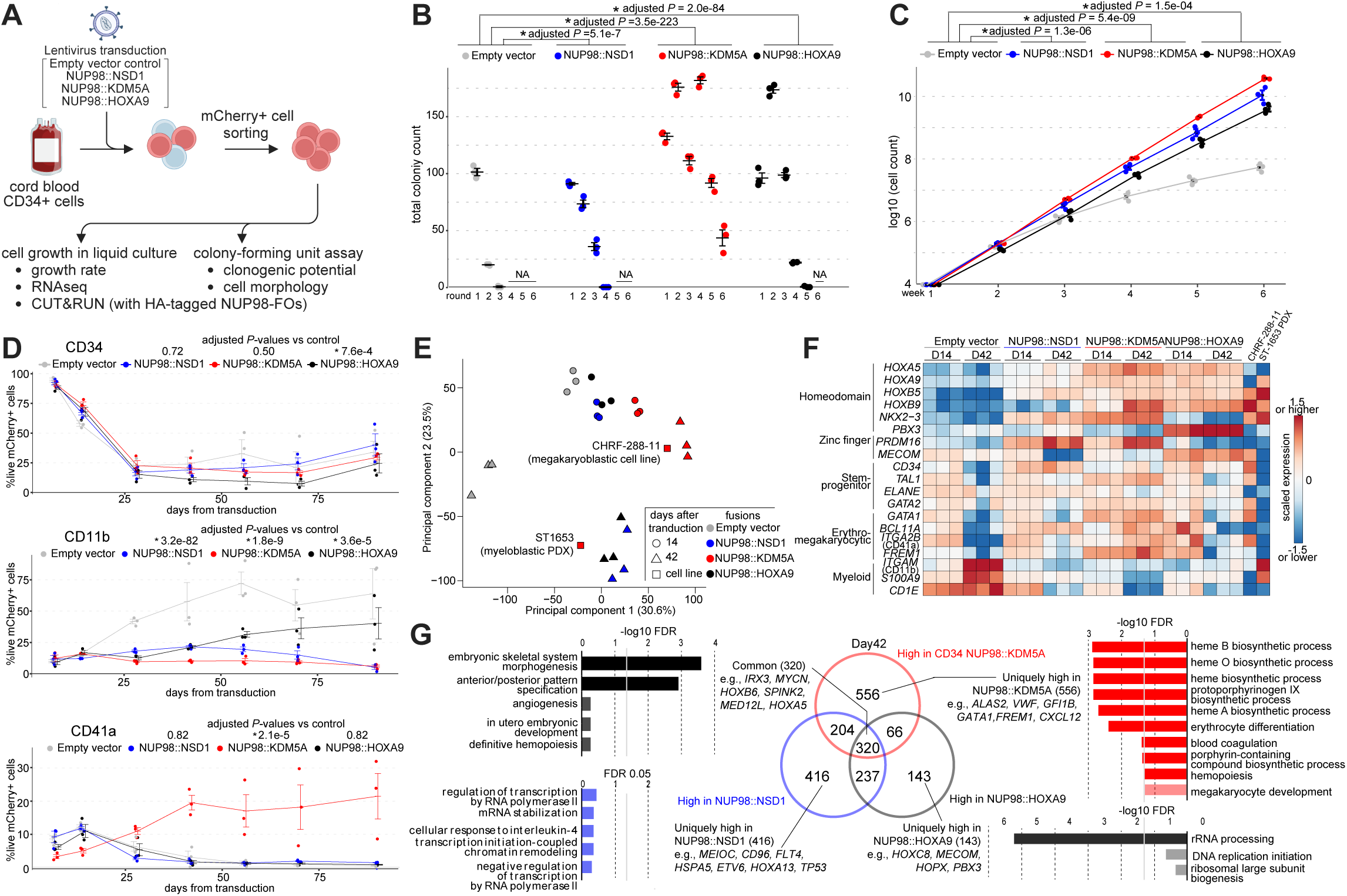
Cord blood CD34 models recapitulate phenotypes of NUP98r leukemia. **A**. Experimental schema using cord blood CD34+ cell models (cbCD34). **B**. Colony-forming unit assays of cbCD34 models with empty control vectors or *NUP98::NSD1*, *NUP98::KDM5A*, or *NUP98::HOXA9*-expressing vectors. **C**. Cell growth assays of cbCD34 models in liquid culture. **D**. Flow cytometric analysis of cbCD34 models in liquid culture (**top**: CD34+, **mid**: CD11b+, **bottom**: CD41a+ population ratio:% in mCherry+ live cells). **E**. Principal component analysis (PCA) of RNAseq data from liquid culture. Colors indicate NUP98 fusions, and shapes indicate days after transduction. **F**. Heatmap showing expression of representative genes related to stemness or differentiation of hematopoietic cells. Colors of cells indicate expression levels normalized among samples, and genes are annotated on the left. **G**. Comparison of differentially expressed genes (DEGs) in each cbCD34 model compared with empty vector controls at day 42. Venn diagram showing overlaps of highly expressed genes in each model (**mid**) and GO term analyses of shared or specific DEGs are shown (**left**, **right**). Data was obtained from three biological replicates (different lots of cord blood). In **B**-**D**, statistical tests were performed by generalized linear mixed effect model with Poisson (**B**) and Gaussian (**C**-**D**) distributions followed by comparison with empty vector control and the Benjamini-Hochberg adjustment, asterisks indicating adjusted *P*-values *<0.05. Error bars indicate mean ± s.e.m. Abbreviations. FO: fusion oncoprotein, PDX: patient-derived xenograft, FDR: false-discovery rate.

### Differential gene regulation by NUP98 fusion oncoproteins

To further study how NUP98-FOs drive leukemogenesis, we profiled the genomic binding of N-terminal HA-tagged NUP98-FOs (Fig.S8A), using anti-HA antibody in cbCD34 models along with histone modifications (H3K4me3 and H3K27ac) and menin using CUT&RUN^21^ (Fig.5A-B, Fig.S8B-D). These studies revealed FO-binding peaks enriched in the promoter and intronic regions of protein-coding genes with high reproducibility among biological replicates (Fig.S8C). All three FOs showed binding to the *HOXA-B* cluster genes, whereas the binding patterns along the anterior-posterior axes were unique to each fusion, corresponding to RNA expression, active histone modifications, and menin binding as previously reported using different models^16–19^ (Fig.5A, Fig.S8D). Comparisons of protein-coding genes with FO-binding peaks showed 97 common target genes for all three NUP98-FOs, including genes involved in leukemogenesis and hematopoiesis (e.g., *RUNX1*, *CDK6*). However, each FO also showed specific targets (NUP98::KDM5A: 5180 genes, NUP98::HOXA9: 684 genes, NUP98::NSD1: 85 genes, Fig.5B). Notably, NUP98::KDM5A uniquely showed peaks on *GFI1B* and *MEIS2*, which are megakaryocyte/erythroid differentiation regulators^51,52^, indicating that direct regulation of differentiation-related genes could drive phenotypes of *NUP98::KDM5A* AMKL.

**Figure 5.**
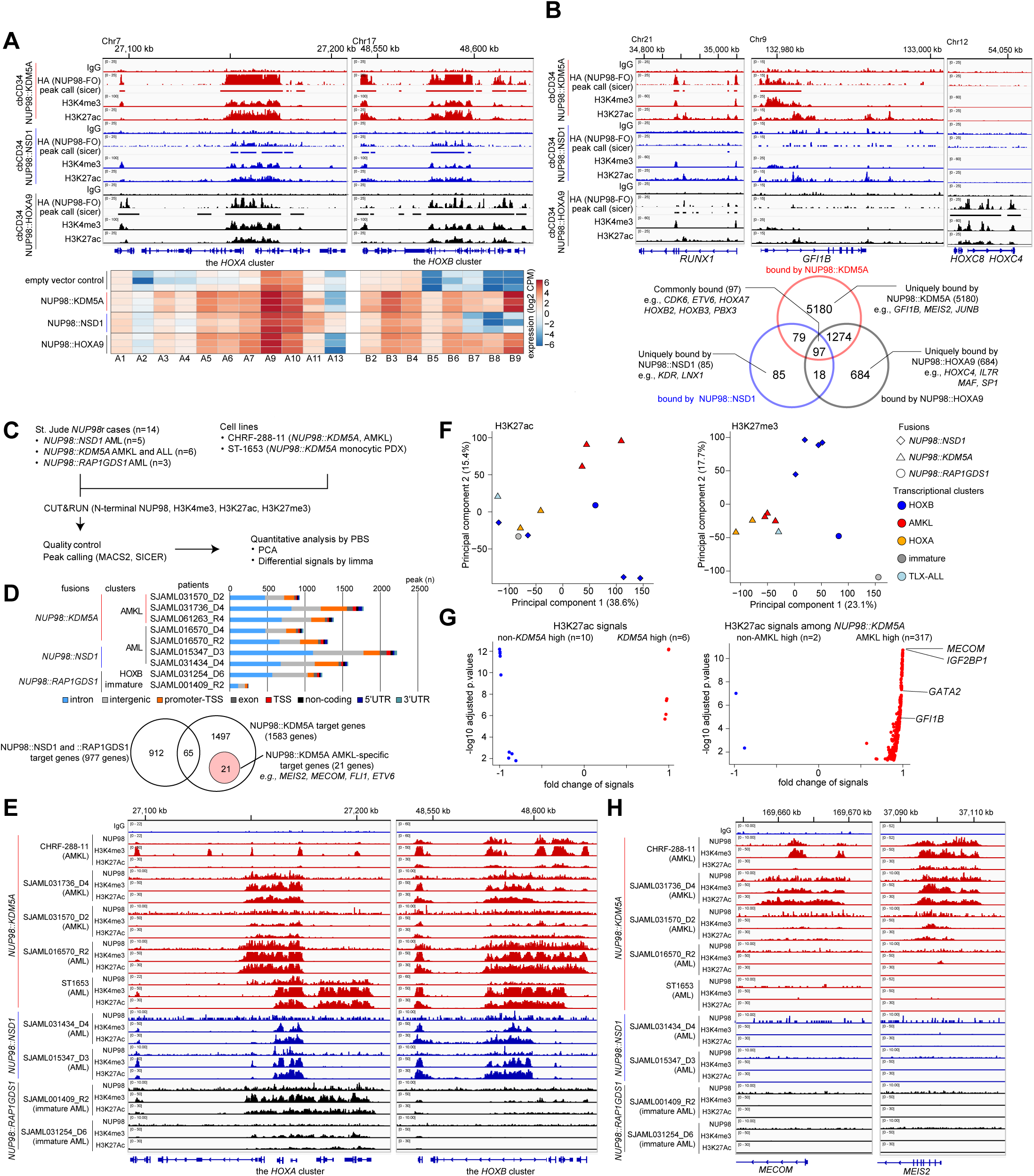
Differential gene regulation by NUP98 fusion oncoproteins. **A**. IGV tracks of the *HOXA-B* clusters from CUT&RUN using HA, H3K4me3, and H3K27ac antibodies in HA-tagged *NUP98*r cbCD34 models (**top**: HA-NUP98::KDM5A-red, HA-NUP98::NSD1-blue, HA-NUP98::HOXA9-black) and heatmap showing expression levels of *HOXA-B* genes (**bottom**) **B**. IGV tracks of differentiation-related gene loci (**top**: *RUNX1*, *GFI1B*, and *MECOM*) and Venn diagram showing overlap of protein-coding genes with annotated peaks (**bottom**: FDR<0.00001). **C**. CUT&RUN strategy from primary patient samples or *NUP98::KDM5A* cell lines (CHRF-288-11 and ST1653). **D**. Counts of peaks from the N-terminus NUP98 antibody in primary samples (**top**) and overlaps of target genes among non-NUP98::KDM5A and NUP98::KDM5A. NUP98::KDM5A AMKL-specific 21 target genes are highlighted. Colors indicate peak annotations. **E**. IGV tracks of the *HOXA-B* cluster from CUT&RUN using N-terminal NUP98, H3K4me3, and H3K27ac antibodies in primary leukemia samples and *NUP98::KDM5A* cell lines. **F**. PCA of genome-wide PBS (probability being signals) scores of H3K27ac (**left**) and H3K27me3 (**right**) from primary samples. Colors indicate expression clusters and shapes indicate fusion partners. **G**. Differential signal analysis using H3K27ac PBS scores between *NUP98::KDM5A* and other (*NUP98::NSD1* and *NUP98::RAP1GDS1*) samples (**left**) and *NUP98::KDM5A* AMKL and non-AMKL (**right**) calculated by limma followed by the Benjamini-Hochberg adjustment. Only regions with significant enrichment (adjusted *P*-values < 0.05) are shown. **H**. IGV tracks of the *MECOM* and *MEIS2* gene loci from CUT&RUN using N-terminal NUP98, H3K4me3, and H3K27ac antibodies in primary leukemia samples and *NUP98::KDM5A* cell lines.

We next sought to profile endogenous NUP98-FO binding without epitope tags in patient samples or cell lines. We tested antibodies targeting the N-terminus of NUP98 in cbCD34, which showed similar binding patterns albeit with weaker signals yielding fewer peaks than HA-antibody, possibly underestimating weak FO-binding (Fig.S9A). N-terminus NUP98 antibody for CHRF-288-11 and ST1653 revealed NUP98 signals at genes highly expressed in these cell lines, such as *HOXA/B* genes and *MEIS1*, with menin co-localization (Fig.S9B). Contrarily, weak or low signals were observed in *RUNX1::RUNX1T1* AML (lacking *HOX* gene expression) or *UBTF-*TD AML (similar to *NUP98::NSD1* AML)^6^. We then applied CUT&RUN using antibodies for N-terminus NUP98 and histone modifications (H3K4me3, H3K27ac, H3K27me3) for 14 *NUP98*r patient samples covering *NUP98::NSD1*, *NUP98::KDM5A*, and *NUP98::RAP1GDS1* (Fig.5C, Fig.S9C, Supplementary Table 12). N-NUP98 antibodies revealed target genes for each FO, including *HOXA* or *HOXB* clusters with heterogeneous patterns even within the same NUP98-FO groups (Fig.5D-E). Also, N-NUP98 antibodies showed NUP98::KDM5A-specific targets (1,518 genes), whereas 21 genes, including *MEIS2* or *MECOM,* were *NUP98::KDM5A* AMKL-specific targets. Genes with H3K4me3 peaks showed a broad overlap among samples, whereas 61 genes, including *IGF2BP3* associated with megakaryocyte development^53^, were unique to *NUP98::KDM5A* AMKL (Fig.S9D). For H3K27ac and H3K27me3 signals with broad peaks that could span multiple genes, we applied the PBS^54^ (probability of being signal) approach to quantify the genome-wide signals, which revealed that genome-wide H3K27ac status correlated with disease phenotypes, whereas H3K27me3 status was associated with fusion partners (Fig.5F). Comparisons of *NUP98::KDM5A* with the other fusions (*NUP98*::*NSD1* and *NUP98*::*RAP1GDS1*) showed limited regions with differential H3K27ac signals (Fig.5G). In contrast, comparison between *NUP98*::*KDM5A* AML and AMKL showed enrichment of *MECOM*, *IGF2BP1*, or *GATA2* regions in *NUP98::KDM5A* AMKL, indicating phenotype-specific epigenetic status (Fig.5G-H, Fig.S9E). These data suggest both the C-terminus of FOs and differentiation status can affect FO-binding patterns to establish heterogenous epigenetic and expression profiles in *NUP98*r leukemia.

### Functional characterization of recurrent somatic alterations in NUP98r leukemia

Given the differential NUP98-FO binding patterns and cooperating mutations related to disease phenotypes, we investigated the impact of co-occurring alterations on *NUP98*r cbCD34 models. We first established cbCD34 models with constitutive Cas9 expression followed by lentiviral transduction of gRNA to mimic *WT1* frameshift mutations or *RB1* loss (Fig.6A, Fig.S10A-B). In NUP98::KDM5A cbCD34 models, *WT1* gRNAs showed enhanced cell growth compared with AAVS-targeting controls, whereas out-of-frame indels in *RB1* were enriched at later time points, indicating growth advantages of cells with *RB1* loss (Fig.6B-C). Morphologically, AAVS-controls showed cells with various degrees of myeloid and erythrocyte/megakaryocyte differentiation, whereas *RB1*-gRNA conditions yielded larger cells with features of maturing megakaryocytes, and *WT1*-gRNA enriched homogenously small blast-like cells (Fig.6B). Flow cytometric analysis showed that *RB1*-gRNA conditions enriched CD34+CD41a+ populations, whereas *WT1*-gRNA conditions depleted the CD41a+ population with increased CD34+CD41a-cells (Fig.6D, Fig.S10C). *NUP98::NSD1* cbCD34 models showed growth advantages with *WT1*-gRNA, whereas *RB1-*gRNAs did not show either growth advantage or consistent enrichment of out-of-frame indels, suggesting that *RB1* loss specifically synergizes with NUP98::KDM5A (Fig.S10D-E). RNAseq revealed global transcriptional changes in *RB1-* or *WT1*-gRNA conditions (Fig.6E). *RB1*-gRNA conditions showed upregulation of *CD34* and *CDKN2A*, which is downstream RB1/E2F and forms a feedback loop^55,56^, and changes in immunity genes compared with AAVS-controls (Fig.6F, Fig.S11A). *WT1*-gRNA also showed increased *CD34* expression, along with increased MHC-class II and decreased megakaryocyte/platelet-related gene expression (Fig.6G).

**Figure 6.**
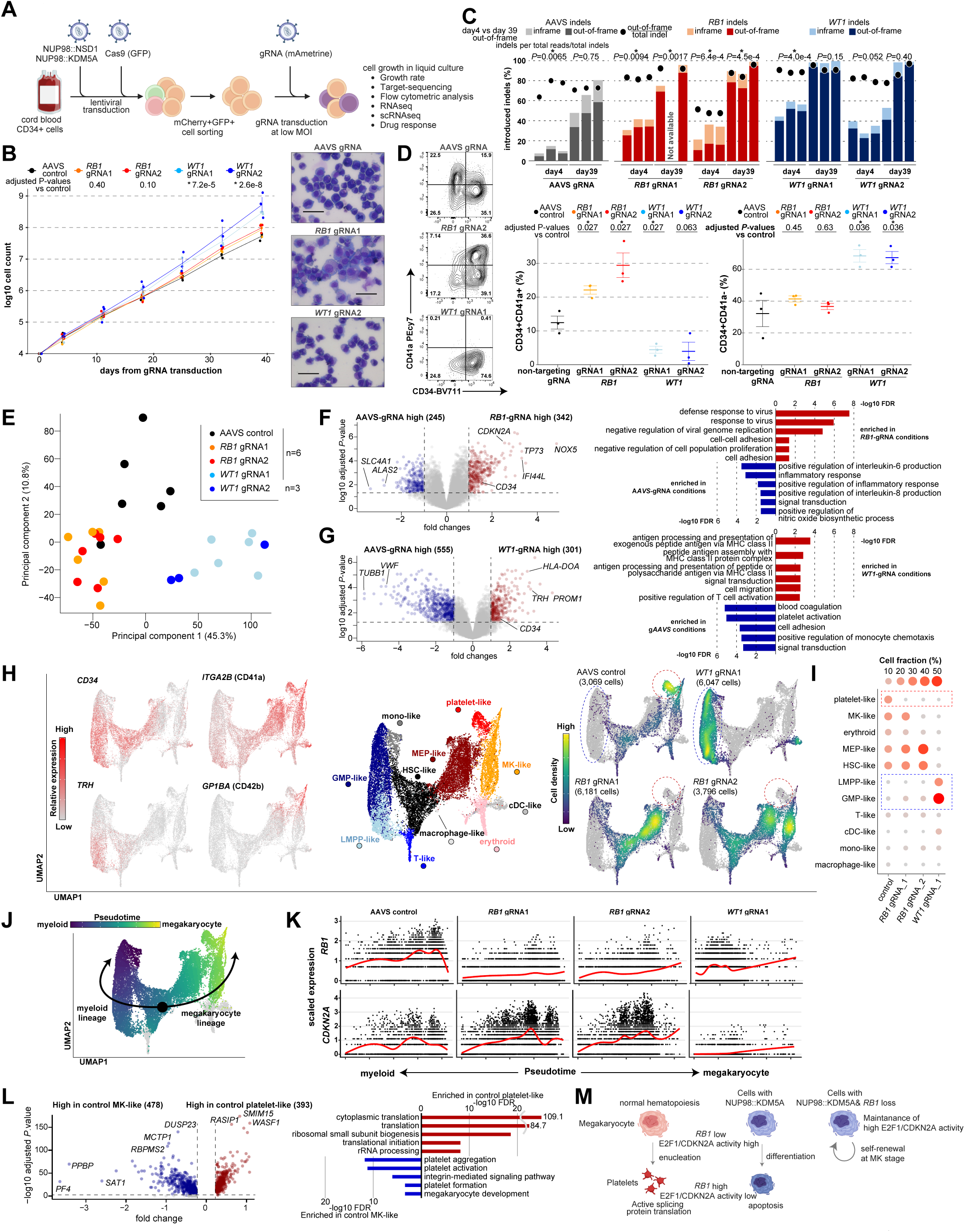
Functional characterization of recurrent somatic alterations in NUP98r leukemia. **A**. Experimental schema of induction of cooperating alterations (*RB1*, *WT1*) in cbCD34/Cas9 models. **B**. Cell growth assays of cbCD34/Cas9 NUP98::KDM5A with gRNAs targeting the AAVS, *RB1*, or *WT1* loci (**left**), and cytospin of cells on day 35 (**right**). **C**. Induction rates of indel (insertions and deletions) at day 4 and 39 in each condition. Bars represent fractions of indel rates in all target sequence reads, and dots represent out-of-frame indel ratio among total indels. **D**. Flow gating (**left**), CD34+ CD41a+ positivity (**mid**), and CD34+ CD41a- (**right**) among mCherry+ GFP+ mAmetrine+ live cells. **E**. PCA of RNAseq of gRNA-transduced NUP98::KDM5A cbCD34 models at day 35. **F**. DEG analysis between AAVS controls and *RB1*-gRNA conditions (**left**) and gene ontology (GO) term analysis of DEGs (**right**). Colors indicate DEGs and GO terms (red: high in *RB1*-gRNA conditions, blue: low in *RB1*-gRNA conditions)**. G**. DEG analysis between AAVS controls and *WT1*-gRNA conditions as shown in **F**. **H**. UMAP plots of scRNAseq data from gRNA-transduced NUP98::KDM5A cbCD34 models at day35, showing marker gene expression (**left**), annotated clusters (**mid**), and cell distributions among conditions (**right**). Colors in plots indicate relative expression levels, clusters, and cell density, respectively. **I**. Enrichment of cells with each cluster indicated by colors and sizes. **J**. Pseudotime along myeloid (HSC→GMP→monocytes) and platelet (HSC→MEP→MK→platelet) trajectories. Colors represent pseudotime scores of each single cell inferred by Slingshot. **K**. *RB1* (**top**) and *CDKN2A* (**bottom**) expression along the pseudotime axis in each condition with red curves show average expressions. **L**. DEG analysis between the platelet-like and MK-like clusters in the AAVS-control condition (**left**) and GO term analysis (**right**) of genes high in the platelet-like cluster (red) and the MK-like cluster (blue). **M.** Schematics illustrating platelet differentiation in normal hematopoiesis and NUP98::KDM5A models. Assay data was obtained in technical triplicates from an established NUP98::KDM5A/Cas9 line and independent experiments. One data point in **C** was not obtained due to technical errors. RNAseq data was obtained from six independent experiments. In **B-D**, statistical tests were performed by linear mixed effect model (**B**) or two-sided Student’s t-test by comparing day 4 and day 39 (**C**) or gRNA conditions and AAVS controls (**D**), and limma (**F**, **G**) followed by the Benjamini-Hochberg adjustment when applicable. DEGs in scRNAseq (**L**) were identified using FindMarker function in Seurat package with default settings, which calculate adjusted *P*-values with limma implementation of the Wilcoxon rank-sum test followed by Bonferroni correction. Asterisks indicating *P*-values or adjusted *P*-values <0.05. Error bars indicate mean ± s.e.m.

To study how co-alterations affected cellular hierarchies, we performed scRNAseq from these conditions, which collectively showed differentiation trajectories resembling normal hematopoiesis (Fig.6H-I, Fig.S11B). A control condition showed broad distributions with enrichment at the platelet-like populations adjacent to the MEP-like or megakaryocyte (MK)-like clusters. By contrast, *RB1*-gRNA conditions showed enrichment of cells at the MEP or MK-like clusters and lacked platelet-like populations, and *WT1*-gRNA showed strong bias at the GMP- and LMPP-like populations rarely found in control and *RB1-*gRNA conditions. Pseudotime analyses showed that *RB1* was upregulated through the platelet differentiation trajectory and highest in the platelet-like cluster, whereas *CDKN2A* expression was high in the MEP or MK clusters but low or undetectable in the platelet cluster (Fig.6J-K, Fig.S11C). Comparison of platelet-like and MK-clusters showed upregulation of translation-related genes in the platelet-like cluster (Fig.6L), recapitulating changes in translation in normal platelet production from MK or proplatelet^57^. We also observed negative correlation with *RB1* and *CDKN2A* among *NUP98::KDM5A* patient samples (Fig.S11D). These suggest that *RB1* loss inhibits terminal platelet differentiation in the NUP98::KDM5A model, leading to the accumulation of cells at the MK-like or MEP-like stages, similarly to megakaryocytic differentiation block seen in E2F transgenic mice^58^ (Fig.6M). The LMPP population enriched in the *WT1*-gRNA condition was characterized by high MHC-class II gene expression and low cell-division gene expression, and inference of cell-cycles from scRNAseq indicated enrichment of cells at G1-stage in the LMPP cluster and at S/G2M in the GMP cluster (Fig.S11E-G), suggesting unique cellular hierarchy of proliferating GMP-like cells and dormant LMPP-like cells in the *WT1*-gRNA condition.

### Cooperating alterations and differentiation status affect sensitivity to menin inhibition

*NUP98*r leukemia has been reported to be dependent on KMT2A/menin complex^18^, and various menin inhibitors are currently being tested in clinical trials enrolling patients with AML including those with *NUP98*r^59^. Mouse models showed menin inhibition led to myeloid differentiation in *NPM1*-mutant and *NUP98*r leukemia models^18,60–62^, although whether menin is required for megakaryocytic lineages or AMKL is currently unclear. We tested a menin inhibitor (revumenib) in NUP98::KDM5A CD34 models with cooperating alterations to assess the sensitivity and impact on cellular populations (Fig.7A). Treatment of AAVS-control conditions showed a decreased cell growth (14.1±4.6%; mean ± standard error, at day 14 of 1µM revumenib treatment compared with DMSO), whereas *RB1-* or *WT1*-gRNA conditions were less sensitive (*RB1*-gRNA: 64.3±10.3%, *WT1*-gRNA, 56.8±4.0%, Fig.7B). Flow cytometric analysis at day 14 showed significant increases in the CD11b+ population in AAVS-control and *WT1-* gRNA conditions, indicating myeloid differentiation, contrasting to an increase in the CD34+CD41a+ population in *RB1*-gRNA conditions (Fig.7C), suggesting adaptive changes of cellular hierarchies. RNAseq analyses of samples on day 7 of treatment showed increased expression of canonical genes suppressed by NUP98-FOs in all conditions, whereas *CDKN2* family^63^, inflammation, and erythroid/megakaryocyte-related genes were enriched in AAVS compared with *RB1*-gRNA conditions, suggesting differentiation induced by menin inhibition (Fig.7D). *WT1*-gRNA showed immunity-related changes as in AAVS controls but more in activated monocyte/lymphocyte genes (e.g., *CD48*, *MAFB,* Fig.S11H), suggesting partial differentiation from dominant GMP or LMPP clusters. We also tested the sensitivity of ST1653-PDX (myelomonocytic) and CHRF-288-11 (megakaryocytic) cells to menin inhibition, which showed that ST1653 was sensitive to menin inhibition but CHRF-288-11 was not, further confirming the association of differentiation status with menin-dependency (Fig.7E). These collective data indicate that cellular differentiation and cooperating mutations can both influence the dependencies within *NUP98*r leukemias (Fig.7F).

**Figure 7.**
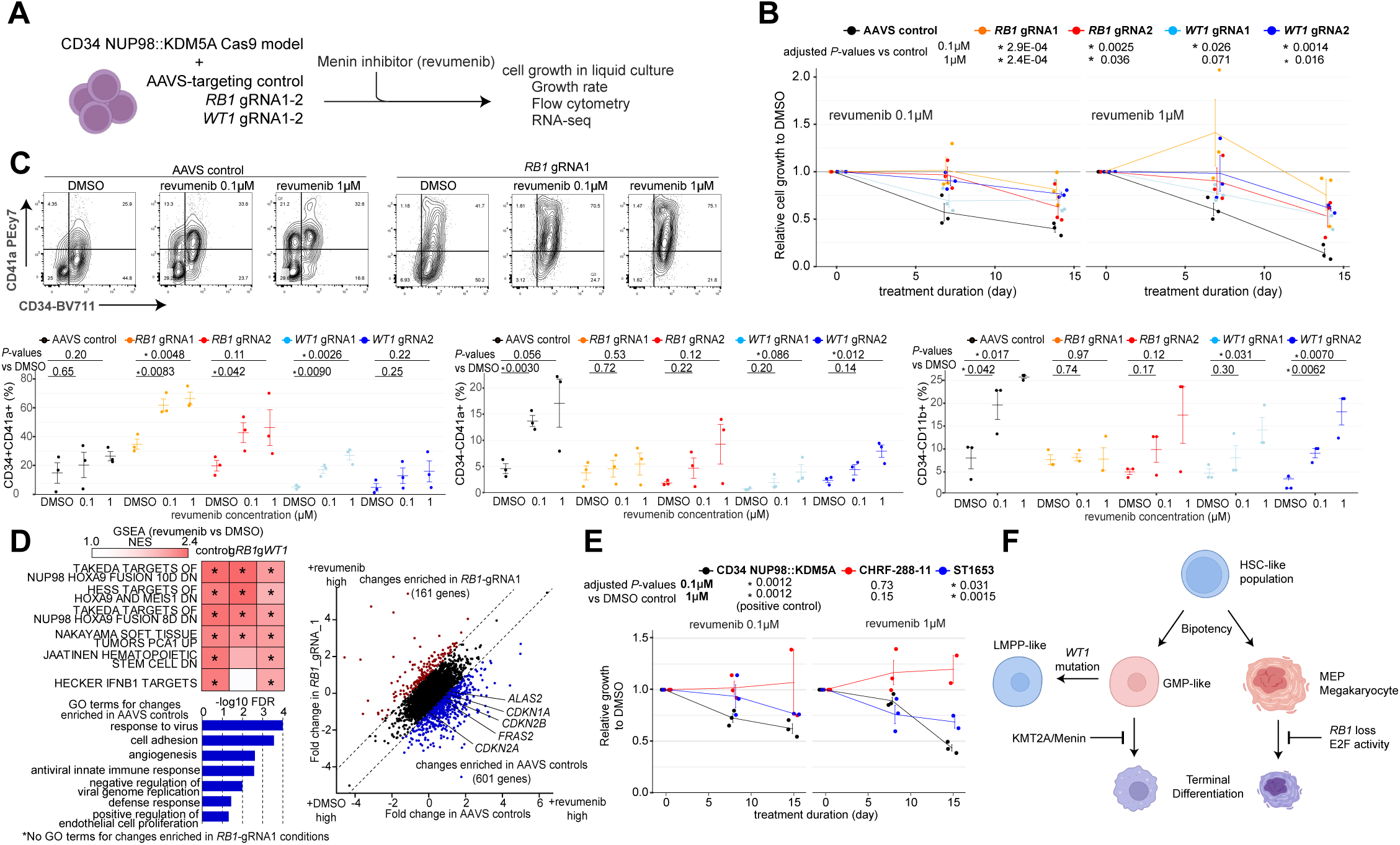
Cooperating alterations and differentiation status affect sensitivity to menin inhibition. **A**. Experimental schema showing revumenib treatment. **B**. Relative cell growth of cbCD34 NUP98::KDM5A models with gRNA treated with revumenib (0.1-1µM) compared with DMSO controls. **C**. Flow gating (**top**), CD34+CD41a+ population (**left**), CD34-CD41a+ population (**mid**), and CD34-CD11B+ populations (**right**) among mCherry+ GFP+ mAmetrine+ live population at day 15. **D.** GSEA analyses between DMSO and revumenib-treated conditions, colors showing NES (**top-left**), comparison of expression changes between AAVS and *RB1*-gRNA1 conditions (**right**), and GO term analysis of changes enriched (difference of fold changes >1) in AAVS conditions (**bottom-left**). **E.** Relative cell growth of unedited cdCD34 NUP98::KDM5A (control, black), CHRF-288-11 (red) and ST1653 PDX (blue) treated with revumenib (0.1-1µM) compared with DMSO controls. **F.** Schematics illustrating cellular hierarchy of NUP98::KDM5A models. Data was obtained from three technical replicates using gRNA-transduced cells in Fig.6. One data point at day 15 was excluded for technical errors. Statistical tests were performed by generalized linear mixed effect model with Gaussian distribution followed by the Benjamini-Hochberg adjustment (**B, E**) or Student’s t-test by comparing gRNA conditions with AAVS controls (**C**). Asterisks indicating *P*-values or adjusted *P*-values *<0.05. Error bars indicate mean ± s.e.m.

## Discussion

Biology-informed treatment of many subtypes of pediatric leukemia is paving the way for improved clinical outcomes^3,64^; however, the biology driving the heterogeneous phenotypes of *NUP98*r leukemia, including somatic alterations or NUP98-FO binding, has not yet been fully characterized. Using genome-wide mutational and transcriptional analyses of 185 *NUP98*r leukemia samples, we expand on the known association of *NUP98*r with *WT1*, *FLT3*-ITD, and *RB1* loss to highlight *JAK2*, *GATA1*, and *MPL* alterations associated with AMKL and *NOTCH1* alterations with T-ALL phenotypes. Profiling with scRNAseq and deconvolution of bulk RNAseq data additionally show heterogeneous cellular hierarchies even with the same fusion partners or in the same patients, suggesting that both fusion partners and cooperating mutations can contribute to the expression profiles and cellular hierarchies^41,43^.

Further, using CUT&RUN in cbCD34 models, we showed that NUP98 FO-binding patterns are unique to each fusion partner, whereas profiling of patient samples and cell lines showed that the binding patterns are not static and could be dynamic according to the differentiation status, represented by AMKL-specific NUP98::KDM5A-binding to *MEIS2* or *MECOM*. Previous knowledge of NUP98-binding patterns is derived from mouse models^16–18^ or in a human non-hematopoietic cell model^19^, whereas our data emphasize the need for studies using human hematopoietic cells to profile NUP98 FO-binding and related epigenetic status contributing to the pathogenesis of *NUP98*r leukemia.

We also show that the introduction of cooperating alterations, namely *RB1* deletion and *WT1* frameshift mutations, could render cells with growth advantages and alter the cellular hierarchies of *NUP98*r models *in vitro* in a fusion partner-specific manner. Previously, transplant models of cbCD34 NUP98::KDM5A cells showed heterogenous disease phenotypes^49^, and whether these cooperating mutations can affect phenotypes *in vivo* is to be investigated in future studies. Although acquired mutations in the *MEN1* gene have been clinically observed as mechanisms of resistance to menin inhibitors in other AML subtypes^65,66^, our experimental models also suggest that cellular differentiation-specific menin dependency can be alternative resistance mechanisms for *NUP98*r leukemia. It will be critical to investigate clinical outcomes of *NUP98*r leukemia treated with menin inhibitors according to fusion partners, cooperating mutations, or expression profiles to identify patients who will not benefit from menin inhibition and treat them with alternative approaches.

Mechanistically, Rb1 is a well-known repressor of E2F1 proteins, which in turn activate the expression of cell cycle-related genes including *CDKN2* families^55,56^. E2F1 activity inhibits normal platelet differentiation leading to the accumulation of abnormal megakaryocytes in E2F1 transgenic mice^58^, whereas *Cdkn2a*-deficient mice showed enhanced platelet production^67^. NUP98-FO binding data from cbCD34 models suggests that preferential differentiation toward MEP and platelets is NUP98::KDM5A-intrinsic, whereas *RB1* loss could lead to high E2F activity and high expression of *CDKN2* family at the MEP/MK stages, which lead to accumulation of megakaryocytic cells and ultimately development of AMKL. However, our cbCD34 models do not fully explain why *NUP98::KDM5A* AMKL is exclusively found in infants and *NUP98::KDM5A* myeloblastic AML is only rarely found in adolescence. Future studies using mice at the developmental stages or transplantation models could dissect the contribution of cell-of-origin (fetal or adult definitive hematopoiesis) or microenvironments (fetal liver or bone marrow) to the *NUP98*r leukemogenesis and provide mechanistic insights needed for better treatment for these refractory diseases.

## Methods

### Subject cohorts and sample details

Tumor samples from patients with acute leukemia from the St. Jude Children’s Research Hospital tissue biorepository were obtained with written informed consent in clinical trials ^68,69^ using a protocol approved by the St. Jude Children’s Research Hospital institutional review board (IRB). Sequence data of tumor samples from the AIEOP (Associazione Italiana Ematologia/Oncologia Pediatrica) and Perugia University were also obtained with written informed consent in each institute. Studies were conducted in accordance with the International Ethical Guidelines for Biomedical Research Involving Human Subjects. No patient received compensation for the enrollment to this study. Patient names were de-identified and replaced with unique research IDs, which are maintained using an honest broker, in accordance with institutional protocols and ethical guidelines. This unique research ID cannot be linked to patient identification outside of the research team. Samples for RNA sequencing (RNAseq: n=10), whole genome sequencing (WGS: n=2), and whole exome sequencing (WES: n=2) were newly sequenced in this study, and the rest of the data were obtained from previous publications (Supplementary Table 1-4) ^3,5,6,8,12,13,22–28^ or public databases (see details in **Data availability** and Table S1). Single cell RNA sequencing data of a *NUP98::NSD1* case was obtained from GEO ^43^ (GSM7494309, GSM7494310).

#### Genotype fingerprints

We performed a pairwise genotype concordance comparison using the estimated genotype from SNPs with ≥20X coverage in RNAseq BAM files to make sure that the study cohort cases represent unique individuals as we performed previously ^5^. We set genotype concordance percentage cutoff ≥90% of SNPs shared between two individuals to identify potential duplicates.

#### Sample processing, library preparation, and sequencing

For newly sequenced samples with low tumor purity (below 60%), the leukemic cell population was enriched either by flow cytometric sorting or T cell depletion by magnetic beads (EasySep Human CD3 Positive Selection Kit II, StemCell Technologies, 17851). For flow cytometric sorting, CD45^dim^CD33^dim^ positive population was sorted using anti-CD45 PerCP-Cyanine5.5 (eBioscience, cat# 8045-9459-120, Clone:2D1, RRID: AB_1907397, 1:20 dilution), anti-CD33 APC (eBioscience, cat# 17-0338-42,clone:WM53, RRID:AB_10667893, 1:20 dilution), and DAPI (BD Biosciences, cat# 564907, RRID:AB_2869624) using FACSAria III instrument and FACS Diva v9.0 (both BD Biosciences) as we have reported ^5^. CD34 gating using anti-CD34 PE (Beckman cat# IM1459U, Clones:QBEnd10; Immu133; Immu409, RRID:AB_131210, 1:5 dilution) was added depending on the positivity of each patient sample. Enrichment of the tumor population was confirmed by flow cytometric analysis of the post-sorting samples (generally > 90%). Libraries were constructed using the TruSeq Stranded Total RNA Kit, with Ribozero Gold (Illumina, 20020598) for RNAseq of patient samples and *NUP98*r cbCD34 models in Fig.4, Ovation RNA sequencing system v2 (Tecan, 7102-32) for the first batch of *NUP98*r CD34/Cas9 models in Fig.6-7, and Illumina Stranded Total RNA Prep, Ligation with Ribo-Zero Plus (Illumina, 20040529) for the second batch. The TruSeq DNA PCR-Free Library Prep Kit (Illumina, 20015963) was used for WGS, and the TruSeq Exome Kit v1 (Illumina, 20020614) for WES according to the manufacturer’s instructions. After library quality and quantity assessment, samples were sequenced on HiSeq2000 or 2500 (Illumina, RRID:SCR_020132, RRID:SCR_016383) instruments with paired-end (2 x 101 bp, 2 x 126 bp, or 2 x 151 bp) sequencing using TruSeq SBS Kit v3-HS (Illumina, FC-401-3001) or TruSeq Rapid SBS Kit (Illumina, FC-402-4023) and HiSeq Control Software with most recent version at the time of sequencing.

#### Whole genome and whole exome sequencing data analysis

The previous genomic lesion calls for the cases (WGS; n=73, WES; n=46) from published studies ^5,6,8,12,13,23–25,27,28^ were collected from their respective publications. For the unpublished cases with DNA data (WGS; n=2, WES; n=2), DNA reads were mapped using BWA ^70,71^(WGS: v0.7.15-r1140 and v0.5.9-r26-dev; WES: v0.5.9-r26-dev and v0.5.9, RRID:SCR_010910) to the GRCh38/hg38 human genome assembly. Aligned files were merged, sorted, and de-duplicated using Picard tools 1.65 (broadinstitute.github.io/picard/). SNVs and Indels were called using Bambino. For cases paired with matched germline controls, germline variants were filtered out if it’s present in the matched germline sample. For unpaired cases, possible germline variants were filtered and classified. Structural variations (SV) were analyzed using CREST^72^ (v1.0, RRID:SCR_005257), and CNVs were analyzed using CONSERTING (1.0) ^73^ on the WGS data. CNVs were also called on cases with only WES DNA data using the following methods. Briefly, Samtools^74^ (v1.16, RRID:SCR_002105) mpileup command was used to generate a mpileup file from matched germline and tumor BAM files with duplicates removed. If a matched germline was not available, a high-quality normal sample was used to pair with the tumor sample. VarScan^75^ (v2.3.5, RRID:SCR_006849) was then used to take the mpileup file to call somatic CNVs after adjusting for normal/tumor sample read coverage depth and GC content. Circular Binary Segmentation algorithm^76^ implemented in the DNAcopy R package (v1.52.0, RRID:SCR_012560) was used to identify the candidate CNVs for each sample. B-allele frequency info was also used to assess allelic imbalance.

#### RNAseq mapping, fusion detection, and large-scale copy number variant calling

RNA reads from newly sequenced samples and from publications were mapped to the GENCODE (RRID: SCR_014966) human genome assembly release 38 gene annotation (GRCh38/hg38) using the StrongARM pipeline ^77^. RNAseq data on hg19 from previous studies were re-mapped onto hg38 using STAR (v.2.7.9a, RRID:SCR_004463). Chimeric fusion detection was carried out using CICERO ^31^ (v0.3.0) on hg19 BAM files and the results were lifted over to hg38 using LiftOver (not versioned, RRID:SCR_018160) from UCSC genome browser (RRID:SCR_005780). For the cases with only RNAseq data, RNAseqCNV ^78^ (v1.2.1) was used to call large-scale copy number variants (CNV) as previously performed ^5^. For possible chromosomal loss involving *RB1*, we manually checked allele frequencies of single nucleotide polymorphisms (SNPs) within the *RB1* gene, around *RB1* genes, and broader regions on chromosome 13 to identify allelic imbalance indicative of loss of heterozygosity of the region (Fig.S3B). Those with allelic imbalance and low *RB1* expression (z-score in the transcriptome cohort < -0.5) were regarded as *RB1* loss (Table S6).

#### Somatic mutation calling from RNAseq

To detect single-nucleotide variants (SNV) and insertions and deletions (Indel) from RNAseq data, we applied our previous approach that showed high sensitivity and specificity on selected genes.^5^ In this study, we focused on a panel of previously screened 87 genes and 25 genes with somatic alterations found in DNA data from this study cohort (Supplementary Table 5). Candidate SNVs/Indels were called by Bambino ^79^ (v1.07, RRID:SCR_005649) or RNAindel ^80,81^ (v3.0.4), annotated by VEP^82^ (v95, RRID:SCR_007931), filtered by excluding variants with gnomAD (v2.1.1, RRID: SCR_014964)^83^ population allele frequency >0.1% as possible germline variants, and in turn, classified for putative pathogenicity with PeCanPie/MedalCeremony^84^ (not versioned). Candidate variants with putative pathogenicity were considered germline or artifacts if present in >5% of the cases. Candidate variants were further filtered if the number of supporting reads was ≤5 or if the variant allele fraction (VAF) was ≤5%.

#### Gene expression data summarization, dimension reduction, and clustering

Reads from aligned BAM files were assigned to genes and counted using RSEM ^85^ (v1.3.0, RRID:SCR_000262) with the GRCh38/hg38 GTF file. Expression data of other T-ALL or AML subtypes from previous studies were obtained in gene x sample matrices ^5,12^ and combined with *NUP98*r other leukemia data to establish RNAseq data of 2,321 samples. For a gene to be considered as expressed, we required that at least 4 samples should have ≥ 20 read counts per million (CPM) reads sequenced. The count data were transformed to log2CPM using Voom^86^ available from R package limma ^87^ (v3.50.3, RRID: SCR_010943). We did not perform batch correction between stranded and unstranded library or data from TARGET or not, as this information confounded with disease subtypes (AML vs ALL). The R package Seurat^88–91^(v4.1.0, RRID:SCR_016341) was used for dimension reduction and sample clustering. Briefly, the top 400 variable protein-coding genes were selected using the “vst” method. The expression data were then scaled and used for PCA (Principal Component Analysis), and the top 25 principal components were used for dimension reduction using UMAP^92,93^ (Uniform Manifold Approximation and Projection, RRID:SCR_018217) (n_neighbors=15 and min_dist=0.3). Samples were clustered using the top 25 principal components by first constructing a K nearest-neighbor graph and then iteratively optimizing the modularity using Louvain algorithm (resolution=0.4). Annotations of each cluster are based on enriched subtypes or known expression profiles. For cord blood CD34+ cell (cbCD34) models, data was processed in the same process, and principal component analysis (PCA) was performed using prcomp function. For combined analyses of patient data and cbCD34 models, shared genes in each data set were used to combine, followed by batch correction between patient and in vitro data using the ComBat (RRID:SCR_010974) method available from R package SVA ^94^(v3.42.0, RRID:SCR_012836). UMAP plot was performed using the top 750 variable genes with the following parameters: dims=1:35, min.dist=0.25, n.neighbors=15L. For cbCD34 NUP98 Cas9 models, batch correction was performed between different library preparations.

Differential expressed gene (DEG) analyses of bulk samples were performed by limma^87^, and we set Log2 CPM = 0 if it is < 0 based on the Log2 CPM data distribution. *P*-values were adjusted by the Benjamini-Hochberg method to calculate the adjusted *P*-values using R function p.adjust. Genes with absolute fold change > 1 and adjusted *P*-values < 0.05 were regarded as significantly differentially expressed. Gene Set Enrichment Analysis (GSEA)^95^ was performed by GSEA (v4.2.3, RRID: SCR_003199) using MSigDB gene sets c2.all and c5 (v7.2.1, RRID:SCR_016863), comparing samples or conditions indicated in figure or figure legends. Permutations were done 1000 times among gene sets with sizes between 15 and 1500 genes. Normalized enrichment scores (NES) and FDR for top representative gene sets related to hematopoiesis, leukemia phenotype, biological processes were shown, and all data is available in source data. Gene Ontology (GO) term analyses of DEGs were performed with DAVID^96^ (v6.8, RRID:SCR_001881) using differentially expressed gene, and results for GO term, biological process (GOTERM_BP_DIRECT) were exported.

#### Single cell RNA sequencing (scRNAseq)

ScRNAseq was performed on 8 patients (9 samples) and 4 conditions of cbCD34/Cas9 models. Frozen patient samples were thawed followed by dead cell removal using EasySep Dead Cell Removal Kit (StemCell Technologies, 17899) and resuspended in DPBS (Dulbecco’s Phosphate Buffered Saline,Gibco,14190144)+0.04%BSA (Bovine Serum Albumin, Sigma-Aldrich, A7030-50G). Cultured cbCD34 cells are directly suspended in DPBS+0.04% BSA. Cells were prepared and quantified following the 10x Genomics cell thawing protocol for single cell assays (CG000447, rev B). Cells were isolated for single-cell barcoding and 3’ (*NUP98::NSD1* diagnosis and relapse pair) and 5’ (*NUP98::KDM5A* and *NUP98::RAP1GDS1,* CD34 models) GEX library preparation using the 10x Genomics Chromium Controller Genetic Analyzer (RRID:SCR_019326) and Next-GEM Single Cell V(D)J Reagent Kit (PN-1000128, PN-1000167) following the manufacturer’s protocol. Libraries were pooled and sequenced on an Illumina NovaSeq 6000 (RRID:SCR_016387) using SP Reagent kit v1.5 (Illumina, 20028401). The feature-barcode count data matrix for each sample was generated from raw sequence FASTQ files using Cell Ranger (v3.0, RRID:SCR_017344) and the GRCh38/hg38 human genome assembly. Public scRNAseq data (GSM7494309, GSM7494310) was downloaded as processed count matrix, gene barcodes, and genes, and processed equally with other samples. The output data from Cell Ranger was further analyzed in R environment (v4.0.2, RRID: SCR_001905) using R package Seurat (v4.1.0, RRID: SCR_007322).

For patient data, count data matrices from each patient were combined using the merge function, followed by excluding cells with less than 200 detected genes or greater than 20% mitochondrial transcripts, which was adjusted for the variety of primary cells with low quality, and we further filtered genes to include those in reference bone marrow and thymocyte data set. Feature counts for each cell were normalized to 1 × 10^4^ counts/cell and natural log transformed using the Normalize Data function. The dimensional reduction was performed by principal component analysis (PCA) using the top 1500 variable genes identified by vst analysis using the Find Variable Features function, followed by Jack Straw analysis and UMAP (uniform manifold approximation and projection, RRID: SCR_018217) using the following parameters (dims=1:20, n.neighbors=15L, min.dist=0.5). Clusters were identified by the Find Neighbors and FindClusters functions (resolution = 0.1) and annotated by enrichment of leukemic samples or normal hematopoietic cells. Marker genes were identified by the FindAllMarkers function in Seurat (min.pct = 0.25) and DEGs were identified by FindMarker gene function in Seurat (v4.1.0), both of which calculate adjusted *P*-values with limma implementation of the Wilcoxon rank-sum test followed by Bonferroni correction.

Projection onto the normal bone marrow and thymocyte reference scRNAseq data was performed using MapQuery function in Seurat. Briefly, scaled data were processed through FindTansferAnchors, TransferData, and MapQuery functions, and the reference data and UMAP models were reproduced using following parameters (dims = 1:25, min.dist = 0.5) and used as reference input. Annotations of the cell types are imported from the original publication, and cell density on the original UMAP was calculated by get_density function from ggplot2 library (v3.3.6, RRID:SCR_014601).

Analyses of scRNAseq data of CD34 models were performed by excluding cells with less than 1500 detected genes or greater than 20% mitochondrial transcripts. Regression of cell cycle related gene was applied using CellCycleScoring functions and ScaleData function with the vars.to.regress parameter. The dimensional reduction was performed by PCA using the top 2000 variable genes and UMAP using the following parameters (dims=1:30, n.neighbors=40L, min.dist=0.2). Clusters were identified by the FindNeighbors and FindClusters functions (resolution = 0.35). Cell lineages were obtained by getLineages function in Slingshot (v1.6.1, RRID:SCR_017012), specifying monocyte, macrophage, cDC, erythroid, T-like, and platelet-like clusters as terminal clusters. Pseudotime was calculated by getCurves function (approx_points = 300, thresh = 0.01, stretch = 0.8, allow.breaks = FALSE, shrink = 0.99) by excluding macrophage, cDC, erythroid, and T-like population.

#### Cellular deconvolution using CIBERSORT

Inference of cellular hierarchy by CIBERSORT^45^ (RRID:SCR_016955) was performed by the web interface of CIBERSORTx^97^. We first established signature matrix using the bone marrow and thymocyte reference count data as reference, from which data was subsampled to include 500 cells or less for one cell type (total 49623 → 5500 cells) to accommodate with the data limit in the web application. Mature T-cell types and B-lineage cell types are excluded from the signature matrix, considering our *NUP98*r cohort including T-ALL and AML only, and parameters of signature matrix were set as follows: replicates: 5, sampling: 0.5, fraction: 0, k.max: 999, q.value: 0.01, G.min: 300, G.max: 500. Deconvolution of bulk RNAseq data was performed in absolute mode with S-mode batch correction with 100 permutations using normalized CPM values as input.

#### CUT&RUN (cleavage under targets and release using nuclease)

CUT&RUN from cbCD34 models and primary leukemia samples were performed using CUTANA™ ChIC/CUT&RUN Kit (SKU: 14-1048) according to manufacturer’s instructions. Briefly, 5.0×10^5^ cells per condition were used as a maximum input, whereas the input cells could be as low as 5.0×10^4^ depending on the availability of samples. For primary samples, frozen samples were thawed followed by dead cell removal using EasySep Dead Cell Removal Kit (StemCell Technologies, 17899), generally resulting in 85-100% of viable cells, and by additional nuclear extraction steps according to the protocol version 3, section 3.2, which was critical to detect endogenous NUP98-FO detection. Cells from cbCD34 models were processed without nuclear extraction. Extracted DNA was quantified using a Qubit fluorimeter (Thermo Fisher, RRID:SCR_018095) and dsDNA Quantification Assay Kits, High Sensitivity (Thermo Fisher, Q32854). Antibodies against H3K27ac (Cell Signaling, 8173S, clone: D5E4, RRID:AB_10949503), H3K4me3 (Epicypher, 13-0041, Mixed Monoclonal, RRID:AB_3076423), H3K27me3 (Cell Signaling, 9733, clone: C36B11, RRID:AB_2616029), menin (Bethyl Cat# A300-105A, polyclonal, RRID:AB_2143306), KMT2A (Epicypher, 13-2004, unknown clone, RRID:AB_3076423), N-terminus NUP98 (Abcam Cat# ab50610, clone:2H10, RRID:AB_881769), N-terminus NUP98 (Cell Signaling. 2288, clone: L205, RRID:AB_561204, for preliminary experiment only), HA (Cell Signaling, 3724, clone: C29F4, RRID:AB_1549585), and control IgG antibody (Epicypher Cat# 13-0042, RRID:AB_2923178) were used at 1:50 dilution in all assays.

#### CUT&RUN library preparation and sequencing

DNA from CUT&RUN experiments were subjected for library preparation using NEBNext Ultra™ II DNA Library Prep Kit for Illumina® (E7645) according to a modified protocol based on protocol.io (dx.doi.org/10.17504/protocols.io.bagaibse). Specifically, DNA from CUT&RUN was used as input (6ng or lower) and adaptor concentrations were proportionally to DNA concentrations (e.g. 3.75 pmol for 6 ng DNA, 0.8 pmol for 1 ng DNA) to avoid extra adaptor dimers. DNA purification with AMPure XP beads were performed as one step (after adapter ligation: 80 µL as 1.75x, after PCR: 36 µL as 1.2x) to enrich short fragments. PCR cycles were set at 12 cycles for 6 ng input and increased up to 15 cycles when input DNA concentration was too low and undetectable. Library amount was quantified using Qubit™ dsDNA Quantification Assay Kits (High Sensitivity) and analyzed by Agilent Bioanalyzer (RRID:SCR_018043) using High Sensitivity DNA chip (Agilent, 5067-4626) before sequencing. Libraries were sequenced on NovaSeq 6000 with paired-end (2×100bp) sequencing using SP Reagent kit v1.5 (Illumina, 20028401) targeting 20 million reads.

### CUT&RUN analytic pipeline

To obtain quality reads, raw reads were processed and trimmed with Trim Galore (v0.4.4, RRID:SCR_011847) from cutadapt (RRID:SCR_011841) and FASTQC analysis. A default quality score cutoff of Q20 is used. Quality reads were mapped to the GRCh38/hg38 human genome assembly using bwa (v0.7.12-r1039, RRID:SCR_010910) and converted to a bam file by samtools (v1.2, RRID:SCR_002105). Duplicate reads were marked with biobambam2 (v2.0.87, RRID:SCR_003308). Uniquely mapped and properly paired reads were then extracted with samtools (v1.2, “-F 1804 -q 1”) and sorted by read name using biobambam2 (v2.0.87, RRID:SCR_003308). bedtools^98^ (v2.17.0, RRID:SCR_006646) were used to convert bam file to bedpe file as fragments. Fragments with size shorter than 2000bp were extracted and center 80bp of each fragment were used to generate bigwig track files by UCSC tools (v4, RRID:SCR_005780) and visualized using Integrative Genomics Viewer (IGV, v2.17.4, RRID:SCR_011793). Peak calling was performed using Macs2 (v2.1.1, RRID:SCR_013291) targeting narrow peaks and SICER (v1.1, RRID:SCR_010843) targeting broad peaks with following command lines.

macs2 callpeak -t $ [treatmentBed} -c $[controlBed] -f BEDPE -g hg38 --keep-dup all -n [”output file”] -q [0.05] sicer -t $ [treatmentBed] -c $ [controlBed] -s hg38 -rt 1 -w 200 -f 200 -egf 0.8-6 -g 600 -fdr 0.00001

Peaks called by macs2 or SICER were filtered according to peak scores (>50, corresponding FDR<1.0e-5) and annotated using HOMER (v4.10, RRID:SCR_010881). Low-quality data with high background signal from IgG controls or low assay signals were excluded according to peak call count and manual inspection of bigwig files on IGV, which are listed in Fig.S9. Overlaps of coding genes with peaks were assessed by hypergeometric test (one-sided) using gmp library (v0.6-5), with total number of coding genes in hg38 (19,385 genes) as a denominator. Quantitative analysis of H3K27ac and H3K27me with broad signals from various primary samples, we utilized PBS^54^ (probability of being signal, v02072024) which is a bin-based strategy of quantification of epigenetic data. PBS was performed by default setting, whose output included normalized signals (0 to 1) on 574,152 genomic regions binned at 5kb length, further filtered for regions with non-zero average signals among samples. PCA was performed by prcomp function, and differential analysis was performed by limma followed by the Benjamini-Hochberg adjustment to calculate adjusted *P*-values. PBS 5kb-binned regions are also annotated using HOMER.

#### Construction of lentivirus vectors and virus Production

*NUP98::NSD1* and *NUP98::HOXA9* cDNA were cloned from patient sample into pDonor221 Gateway entry vector (Thermo Fisher, 12536017) and then transferred to Gateway-compatible vectors (CL20-MSCV-ires-GFP and MND-mPGK-mCherry). *NUP98::KDM5A* expression vectors^49^ were kindly provided by Dr. Tanja A. Gruber and cloned into MND-mPGK-mCherry. N-terminal HA tags (YPYDVPDYA) were introduced by amplification during cloning. cDNA encoding Cas9 expressing lentivirus vector (Addgene, FUCas9GFP, Plasmid #85555) and used without cloning. gRNA expressing vector were established by Center for Advanced Genome Engineering (CAGE) in St. Jude Children’s Research Hospital using validated gRNAs and LentiGuide-Ametrine backbone expressing mAmetrine under mPGK promoter as described previously^99^. These lentivirus vectors were co-transfected with packaging vectors (pHDM-G, pCAGGHIVgpco, and pCAG4-RTR2, provided from the St. Jude Vector Laboratory) into 50%–60% confluent HEK293T cells (CVCL_0063, ATCC #CRL-3216, obtained from ATCC in 2016) using FuGene HD Transfection Reagent (Promega, E2311). Supernatant containing lentiviral particles was harvested at 48 hours after transfection and concentrated for cord blood CD34+ transduction using Amicon Ultra-0.5 Centrifugal Filter Units (MilliporeSigma, UFC905096). HEK293T cells used for virus production were maintained in DMEM (Gibco, #11965) supplemented with 10% Fetal Bovine Serum (FBS, R&D Systems, S11550H) and 100 U/mL penicillin–streptomycin (Gibco, 15140122), which were validated by short tandem repeat (STR) analysis and were generally used within 15 passages after thawing. Mycoplasma testing was intermittently performed using MycoAlert Mycoplasma Detection Kit (Lonza, #LT08–118).

#### Virus transduction of NUP98-FOs into cord blood CD34+ cells

Commercially available cord blood CD34+ cells were purchased from Lonza (catalog no. 2C-101) or the Carolinas Cord Blood Bank/Duke University. After thawing, cells were cultured for 24 hours in StemSpan SFEM II media (STEMCELL Technologies, #09655) supplemented with penicillin–streptomycin, l-glutamine, and recombinant human SCF, FLT-3, TPO, and IL6 (all 50 ng/mL, PeproTech, HHSC6), SR-1 (1 μmol/L, STEMCELL Technologies, 72344), and UM171 (35 nmol/L, STEMCELL Technologies, 72914, for Fig.4) or UM729 (10 nmol/L, STEMCELL Technologies, 72332, for Fig.6-7). Cells were transduced with MND-PGK-mCherry lentivirus expressing untagged NUP98-FO (NUP98::KDM5A, NUP98::NSD1, and NUP98::HOXA9) or empty vector controls at an MOI (multiplicity of infection) of ≥50 by adding 100µL of concentrated virus to 2.0×10^4^ cells in 100µL media (1:1 ratio) supplemented with Polybrene (1µg/ml, Millipore #TR-1003-G) in a 96-well dish (Corning, 3799). Transduced mCherry+ cells were enriched by flow cytometric sorting on day 7 of transduction and plated for colony-forming unit (CFU) assay in MethoCult H4435 (STEMCELL, 04435) at 1.0×10^3^ cells/dish, followed by weekly serial replating. Cell growth assay was performed by weekly passaging cells at the density of 1.0×10^4^ cells in 100 µL/well in 96-well dishes. HA-tagged NUP98-FOs were transduced similarly for CUT&RUN experiments. Experiments were performed in biological triplicates using different lots of cord blood.

#### Virus transduction of Cas9 and gRNA into cbCD34 models

Established cbCD34 models initially generated by transducing NUP98-FO were maintained in supplemented with penicillin–streptomycin, l-glutamine, and recombinant human SCF, FLT-3, TPO, and IL6, SR-1 and UM729 as described above. Cells were then transduced with MND-PGK-GFP lentivirus expressing Cas9 at an MOI of ≥50 by adding 100 µL of concentrated virus to 5.0×10^4^ cells in 100 µL media (1:1 ratio). Transduced mCherry+GFP+ double positive cells were enriched by flow cytometric sorting on day 7 or later and expanded. Transduction of gRNA expressing vectors (AAVS gRNA controls or gRNAs for *WT1* and *RB1*) were performed around day 40-60 from transduction of HA-tagged NUP98-FO, by adding 25µL of concentrated virus to 2.0×10^5^ cells in 100µL media (1:4 ratio) to achieve a low MOI around 0.5. Cell growth assay was performed by weekly passaging cells at the density of 2.0×10^4^ cells/100 µL/well in 96-well dishes (Corning). Confirmation of introduction of indels was performed by target sequencing using genomic DNA from cells at day 39 (cbCD34 NUP98::KDM5A) or day 32 (cbCD34 NUP98::NSD1). Drug sensitivity tests were performed using HA-tagged NUP98::KDM5A/Cas9/gRNA cbCD34 models at day 35 or later from gRNA transduction in 96-well dishes using revumenib (or SNDX-5613, MedChemExpress, cat# HY-136175). Experiments were performed in technical triplicates using established Cas9+ cell lines.

#### Cytospin

For assessment of cell morphology, 100,000 cells were washed with 1X PBS and spun onto Superfrost Plus Microscope slides (Fisher Scientific, 12-660-16) at 800 rpm for 5 min and then subjected to standard Wright-Geimsa staining as previously. Pictures were taken using OLYMPUS BX60 and INFINITY ANALYZE 7 software (TELEDYNE), and TIFF files were resized by Windows Paint.

#### Cell line culture

CHRF-288-11 cell line^46^ (CVCL_A280), which is acute megakaryocytic leukemia cell line with *NUP98::KDM5A*, was kindly provided by Dr. Tanja A. Gruber and maintained in RPMI 1640 (Gibco, 11875093) supplemented with 10% FBS and 100 U/mL penicillin–streptomycin, which were validated by short tandem repeat (STR) analysis. ST1653^50^, which is a patient-derived xenograft (PDX) cell line, was provided by XenoSTART/ The START Center for Cancer Research with a written agreement for use for research purposes and maintained in RPMI 1640 supplemented with 10% FBS and 100 U/mL penicillin–streptomycin. For drug sensitivity test, cells were plated at the density of 2.5×10^5^ cells/ 330 µl in a 24-well dish (Corning, 3524). Mycoplasma testing was intermittently performed using MycoAlert Mycoplasma Detection Kit (Lonza, #LT08–118).

#### Flow cytometric analysis and cell sorting

Cells were washed with FACS buffer (DPBS supplemented 2%FBS and penicillin-streptomycin) and stained with the following combination of antibodies and dyes along with fluorescent proteins (mCherry for Fig.4 and Fig.S7, and mCherry/GFP/mAmetrine for Fig.6-7) Fig.4 and Fig.S7; LIVE/DEAD Fixable Aqua Dead Cell Stain Kit, for 405 nm excitation (Thermo Fisher, L34966), anti-CD34 AlexaFluoro700 (BioLegend, 343526, RRID:AB_2561495, clone: 581, 1:50 dilution), anti-CD11b APC-Cy7 (BioLegend, 301342, clone:ICRF44, RRID:AB_2563395, 1:50 dilution), and anti-CD41a BV605 (BD Horizon™, 563250, clone: HIP8, RRID:AB_2738096, 1:50 dilution). Cells were also stained with anti-CD45 PerCP-Cy5.5 (BD Pharmingen™, 564105, clone: HI30, RRID:AB_2744405, 1:50 dilution), anti-CD33 APC (Invitrogen, 17-0338-42, clone:WM-53, RRID:AB_10667893, 1:50 dilution), anti-CD235a BV421 (BD Horizon™, 562938, clone: GA-R2, RRID:AB_2721016, 1:50 dilution), and anti-CD71 BV711 (BD Horizon™, 563767, clone: M-A712, RRID:AB_2738413, 1:50 dilution) whereas we did not include these signals in the gating strategy in this manuscript.

Fig.6-7 and Fig.S10; anti-CD34 BV711 (BD, 745543, clone:8G12, RRID:AB_2743070, 1:40 dilution), anti-CD11b APC (BD, 561015, clone:ICRF44, RRID:AB_10561676, 1:20 dilution), and anti-CD41 PEcy7 (Biolegend, 303718, clone:HIP8, RRID:AB_10899413, 1:100 dilution).

Cells are incubated at room temperature or on ice for 15-30 min, washed by FACS buffer, and resuspended in FACS buffer with DAPI Staining Solution (Miltenyi Biotec.,130-111-570, 1:1000 dilution, for Fig.6-7 only), and analyzed by BD LSRFortessa X-20 Cell Analyzer (RRID:SCR_025285, Fig.4) or BD FACSymphony A5 Cell Analyzer (RRID:SCR_022538, Fig.6-7). Output FCS files were analyzed FlowJo (v10.10.0, RRID:SCR_008520).

Cell sorting of fluorescent+ (mCherry for NUP98 FOs, and GFP for Cas9) was performed by BD FACSAria III Cell Sorter (RRID:SCR_016695) or Sony MA900 Multi-Application Cell Sorter (no RRID) without staining of cell surface markers.

#### Western blotting

HA-tagged NUP98::KDM5A/Cas9/gRNA cbCD34 models at day 35 or later time point were subjected to Western blotting for RB1 and WT1 proteins. 25 micrograms of purified proteins were separated by SDS-PAGE on a 4-20% gradient gel (Bio-Rad, #4561033) and transferred onto nitrocellulose membranes (0.2 umol/L, Bio-Rad, 1620252). Membranes were incubated overnight at 4°C with mouse anti-Rb (Cell signaling, 9309T, clone: 4H1, RRID:AB_823629 1:1000 dilution), rabbit anti-WT1 (Cell signaling, 83535, clone: D8I7F, RRID:AB_2800020, 1:1000 dilution) or anti-GAPDH (Cell signaling, 2118S, clone 14C10, RRID:AB_561053, 1:1000 dilution). Amersham ECL Rabbit IgG, HRP-linked F(ab’)₂ fragment (from donkey)(Cytiva, NA9340, 1:2,000 dilution, RRID:AB_772191) and Sheep Anti-Mouse IgG-Horseradish Peroxidase (Cytiva, NA931, 1:2,000 dilution, RRID:AB_772210) were used as secondary antibodies. Imaging was performed on the BIO-RAD Chemidoc Touch imaging system and processed using Image Lab (v6.1).

#### Statistics & Reproducibility

No sample size, power calculation, or randomization of patients was performed in this study utilizing retrospective profiling of patients with available materials or sequence data. No analysis depending on patient background was performed in this study. No blinding was performed in the enrollment of patients or data collection of public data, and blinding in group allocation was not possible as the grouping is based on the molecular characteristics of individual patients. For discrete values of the molecular category and the mutation frequency in cohorts, statistical significance and mutual exclusivity were assessed by two-sided Fisher’s exact test using fisher.test function. Comparison of variant allele frequencies (VAFs) or CIBERSORT scores were performed by the two-sided Wilcoxon rank-sum test using wilcox.test function. Adjustment of multiple testing was performed by the Benjamini-Hochberg method using p.adjust function on R when appropriate. The effect size for Pearson’s correlation and statistical significance were assessed by cor and cor.test functions on R.

Experiments using cord blood cells are performed in biological triplicates using cord blood cells from different donors (Lonza, cat# 2C-101A,, Donor#24690, Donor#25195, Donor#24987). Experiments of cooperating mutations using CRISPR/Cas9 system were performed using cbCD34 model lines with Cas9, one for each NUP98::KDM5A and NUP98::NSD1, and in technical triplicates from three independent experiments at different passages and one week apart. Sample size (n=3) was determined according to preliminary experiment. Statistical significance was assessed using generalized linear mixed effect model for growth assays and colony assays using glmer (lmer) function (lme4 package, v1.1-35.1) and emmeans function (emmeans package, v1.5.0), and unpaired two-sided Student’s t-test with equal variances (t.test function) assuming normality according to preliminary experiments, followed by adjustment of multiple testing was performed by the Benjamini-Hochberg method using p.adjust function on R when appropriate. Missing data was treated as null in statistical analyses and indicated in figures or legends. Definition of box plots or error bars are included in figure legends of each figure, and individual data points were shown when possible. Data used for figures are included in source data spreadsheets.

#### Visualization

Mutational heatmaps, pie charts, and bar plots were visualized using Windows Excel. Schemas for fusion partners were made by ProteinPaint (https://proteinpaint.stjude.org/). Heatmaps of expression data and mutual-exclusivity matrix were made by the pheatmap library (v1.0.12, RRID:SCR_016418). Other data visualizations were performed by the ggplot function of the ggplot2 library (v3.3.6, RRID:SCR_014601), the ggbeeswarm library (v0.7.2), Seurat package (v4.0.2, RRID:SCR_016341) and the base plot function in the R statistical environment (v4.0.2). Schemas were created with BioRender.com (RRID:SCR_018361). Figures are incorporated and edited using Adobe Illustrator (2021, RRID:SCR_010279).

## Data availability

Genomic analyses in this study were based on the GENCODE GRCh38/hg38, and gnomAD (v2.1.1, RRID: SCR_014964) was used for classification for germline and somatic mutations. The genomic data and expression data newly generated in this study (RNAseq: n=10, WGS: n=2, WES: n=2) and undeposited data from previous studies (RNAseq: n=3) have been deposited in the European Genome-Phenome Archive (EGA, RRID:SCR_004944), which is hosted by the European Bioinformatics Institute (EBI), under accession EGAS00001005760. For the remaining RNAseq data for 175 samples, 56 samples are sequenced in St. Jude and available either on EGA or St. Jude Cloud ^5,6,8,13,28^ or on the original publication ^26^. For 9 published cases ^27^, we downloaded the BAM files from EGA (EGAS00001004701). Of the remaining WGS data for 73 samples, 34 are sequenced at St. Jude samples, data for all 34 samples from the original publications^5,6,13,28^ are available on either EGA or St. Jude Cloud, and for other 7 published cases ^27^, we downloaded the BAM files from EGA (EGAS00001004701). For the remaining WES data for 46 samples, 21 are sequenced at St. Jude and all data from the original publications ^5,6,8,13,24,28^ are available either on St. Jude Cloud or EGA, and for the other 9 published cases ^27^, we downloaded the BAM files from EGA (EGAS00001004701).

The data generated by the TARGET-AML initiative ^23,25^ (RNA:n=18, WGS:n=18, WES:n=2) is available under accession [phs000218](https://portal.gdc.cancer.gov/projects/TARGET-AML) (TARGET-AML) and [phs000465](https://portal.gdc.cancer.gov/projects/TARGET-AML) (TARGET sub-study, data is available as a part of phs000218), managed by the NCI. Data from the AAML1031 study lead by COG is also available at GDC data portal under [phs000218](https://portal.gdc.cancer.gov/projects/TARGET-AML) (TARGET-AML). Public data from the COG AALL0434 trial (RNA:n=14, WGS:n=14, WES:n=14) is available on dbGaP under phs002276.v2.p1 (phs000218 and phs000464 for T-ALL TARGET samples) and the Kids First data portal (https://portal.kidsfirstdrc.org/dashboard). Information about TARGET can be found at http://ocg.cancer.gov/programs/target. Background RNAseq data of other T-ALL^12^ was also obtained from SYNAPSE under project ID of syn54032669, and data of other AML subtypes were from publication^5^. All public sequencing data and relevant clinical information were obtained with de-identified sample/patient IDs.

Output files from Cell Ranger for scRNAseq data of 9 samples from 8 *NUP98*r patient and 4 samples from cbCD34 models, bulk RNAseq data (log2CPM count data) from cbCD34 models, CUT&RUN data (.bigwig files) from primary patient samples and CD34 models have been deposited to GEO and will be available upon publication. Public patient scRNAdata from the COG^43^ (GSM7494309, 7494310) is available at GEO under accession number GSE235063, and the patient data was obtained from supplemental tables from the publication. Other data generated in this study are available in Supplemental tables or Source data files or upon request to the corresponding author. We did not use any custom code in this study.

## Acknowledgments

We thank all the patients and their families at St. Jude Children’s Research Hospital (SJCRH) for their contribution to the biological specimens used in this study. We also thank the Biorepository, the Flow Cytometry and Cell Sorting Core, the Center for Applied Bioinformatics (CAB), the Center for Advanced Genome Engineering (CAGE), and the Hartwell Center for Biotechnology at SJCRH for their essential services, which are supported by the NIH (P30 CA021765, Cancer Center Support Grant). This work was funded by the American Lebanese and Syrian Associated Charities of St. Jude Children’s Research Hospital and grants from the NIH (U54 CA243124, Fusion Oncoproteins in Childhood Cancers (FusOnC2) Consortium granted for C.G. Mullighan and J.M. Klco, R35CA197695 for C.G. Mullighan, and R01 CA276079 to J.M. Klco). The content, however, does not necessarily represent the official views of the NIH and is solely the responsibility of the authors. J.M. Klco holds a Career Award for Medical Scientists from the Burroughs Wellcome Fund and is a previous recipient of the V Foundation Scholar Award (Pediatric). D.D.G received financial support under the National Recovery and Resilience Plan (NRRP), Mission 4, Component 2, Investment 1.1, Call for tender No. 1409 published on 14.9.2022 by the Italian Ministry of University and Research (MUR), funded by the European Union-NextGenerationEU, Project Title “Experimental modeling to identify molecular mechanisms and therapeutic vulnerabilities in NUP98-rearranged leukemia“, CUP:J53D23017510001-Grant Assignment Decree No.1384 adopted on 01/09/2023 by the Italian Ministry of Ministry of University and Research (MUR).

