## Supplementary Figures 1-11 for "Fusion oncoproteins and cooperating mutations define disease phenotypes in *NUP98*-rearranged leukemia"

### Fig.S1

**A**

| study/trial | The National Clinical Trial (NCT) number | PMID | author |
|---|---|---|---|
| the AAML1031 trial (COG) | NCT01371981 | PMID: 35349331<br>PMID: 36815378 | Pollard, et al<br>Bertrums, et al |
| the pediatric AML classification study | | PMID: 38212634 | Umeda, et al |
| the relapsed pediatric AML study | | PMID: 35176137 | Umeda, et al |
| the AALL0434 trial (COG) | NCT00408005 | PMID: 39143224 | Pölönen, et al |
| the TARGET AML study (COG) | | PMID: 29227476 | Bolouri, et al |
| the pediatric AML novel subtype study | | PMID: 34778799 | Fornerod, et al |
| the TARGET AML-IF (induction failure) study (COG) | | PMID: 30760869 | McNeer, et al |
| the pediatric non-Down syndrome acute megakaryoblastic leukemia study | | PMID: 28112737 | de Rooij, et al |
| the therapy-ralated myeloid neoplasm study | | PMID: 33579957 | Schwartz, et al |
| the acute erythroleukemia study | | PMID: 30926971 | Iacobucci, et al |
| the pediatric myelodysplastic syndrome study | | PMID: 29146900 | Schwartz, et al |
| the Pediatric Cytogenetically Normal FLT3-ITD AML study | | PMID: 31350825 | Buelow, et al |
| the NUP98-PHF23 fusion study (AIEOP) | | PMID: 26066811 | Togni, et al |
| new cases from AIEOP | | | |
| new cases from Perugia University | | | |
| new case from the RELHEM trial | NCT00908167 | PMID: 21768474 | Inaba, et al |
| new case from the SELHEM trial | NCT02212561 | PMID: 27507877 | Alexander, et al |

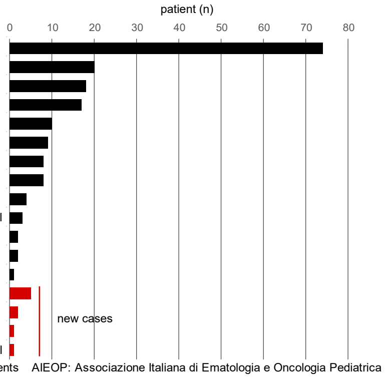


COG: The Children's Oncology Group TARGET: The Therapeutically Applicable Research to Generate Effective Treatments AIEOP: Associazione Italiana di Ematologia e Oncologia Pediatrica

**B**

| Fusions | patient (%) | patient (n) | sample (n) | annotation | *NUP98* exon breakpoint 8 | 10 | 11 | 12 | 13 | 14 | 16 | known domains |
|---|---|---|---|---|---|---|---|---|---|---|---|---|
| *NSD1* | 51.7% | 92 | 98 | epigenetics | 0 | 0 | 0 | 89 | 1 | 0 | 1 | PhD finger/Bromodomain |
| *KDM5A* | 25.3% | 45 | 46 | epigenetics | 0 | 0 | 0 | 1 | 44 | 0 | 0 | PhD finger/Bromodomain |
| *PHF23* | 2.2% | 4 | 4 | epigenetics | 0 | 0 | 0 | 0 | 4 | 0 | 0 | PhD finger/Bromodomain |
| *TNRC18* | 1.1% | 2 | 2 | epigenetics | 0 | 0 | 0 | 0 | 2 | 0 | 0 | BAH1and coiled-coil domain |
| *JADE2* | 1.1% | 2 | 2 | epigenetics | 0 | 0 | 0 | 0 | 1 | 0 | 1 | PhD finger/Bromodomain |
| *PSIP1* | 0.6% | 1 | 1 | epigenetics | 1 | 0 | 0 | 0 | 0 | 0 | 0 | LEDGF domain (four helices) |
| *ZFX* | 0.6% | 1 | 1 | epigenetics | 0 | 0 | 1 | 0 | 0 | 0 | 0 | C2H2 zinc finger domain |
| *BRWD3* | 0.6% | 1 | 1 | epigenetics | 0 | 0 | 0 | 1 | 0 | 0 | 0 | Bromodomain |
| *BPTF* | 0.6% | 1 | 1 | epigenetics | 0 | 0 | 0 | 0 | 1 | 0 | 0 | PhD finger/Bromodomain |
| *HOXD13* | 1.7% | 3 | 3 | homeobox | 0 | 0 | 0 | 2 | 0 | 0 | 1 | homeobox |
| *HOXA13* | 1.1% | 2 | 2 | homeobox | 0 | 0 | 0 | 2 | 0 | 0 | 0 | homeobox |
| *HHEX* | 1.1% | 2 | 2 | homeobox | 0 | 0 | 0 | 2 | 0 | 0 | 0 | homeobox |
| *HOXA9* | 0.6% | 1 | 1 | homeobox | 0 | 0 | 1 | 0 | 0 | 0 | 0 | homeobox |
| *PRRX1* | 0.6% | 1 | 1 | homeobox | 0 | 0 | 1 | 0 | 0 | 0 | 0 | homeobox |
| *RAP1GDS1* | 6.2% | 11 | 12 | signaling | 0 | 1 | 8 | 2 | 0 | 0 | 0 | Armadillo repeat (multiple structured helices) |
| *DDX10* | 1.1% | 2 | 2 | RNA-binding | 0 | 0 | 0 | 0 | 0 | 2 | 0 | srmB domain (three helices) |
| *DDX1* | 0.6% | 1 | 1 | RNA-binding | 0 | 0 | 0 | 0 | 0 | 0 | 1 | srmB domain (three helices) |
| *HMGB3* | 0.6% | 1 | 1 | DNA structure | 0 | 0 | 0 | 0 | 0 | 1 | 0 | HMG domain (four helices) |
| *CEP295* | 0.6% | 1 | 1 | DNA structure | 0 | 0 | 0 | 0 | 1 | 0 | 0 | ALMS domain (three helices) |
| *LNP1* | 1.1% | 2 | 2 | no known function | 0 | 0 | 0 | 0 | 2 | 0 | 0 | no known domain (three helices) |
| *CCDC28A* | 0.6% | 1 | 1 | no known function | 0 | 0 | 0 | 0 | 1 | 0 | 0 | coiled-coil domain |

**C**

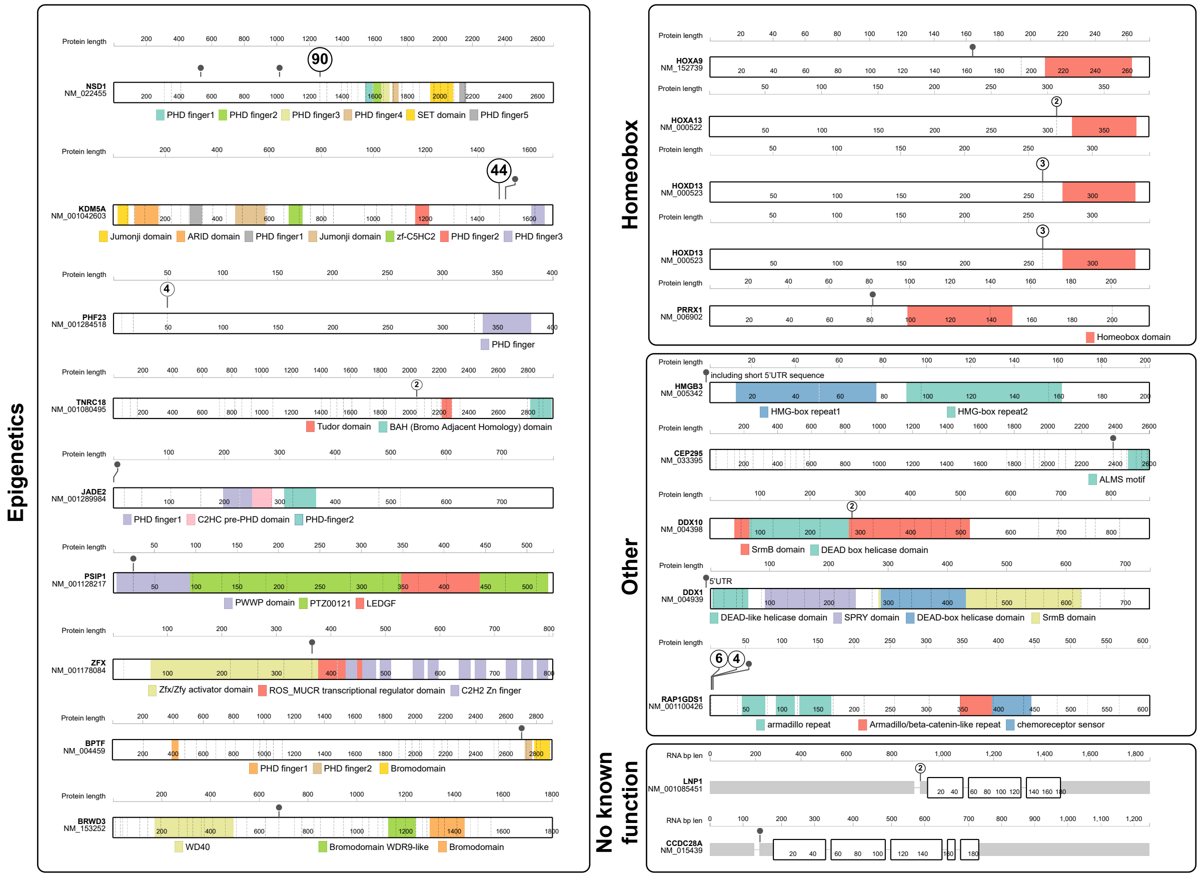


#### Figure S1. *Summary of patients and fusion partners in the NUP98-rearranged leukemia cohort*

**A**. Sources of patient samples (n=185) with publication and trial information when available. Cases newly sequenced for this study (n=10) are colored in red. **B**. Summary tables of fusion partners in the study cohort. Recurrence of *NUP98* breakpoint exons in each fusion partner are colored in red. **C**. ProteinPaint illustrations of fusion partner proteins grouped according to functional annotations. Representative domains are colored as indicated at each figure, and breakpoints and numbers are indicated by circles.

### Fig.S2

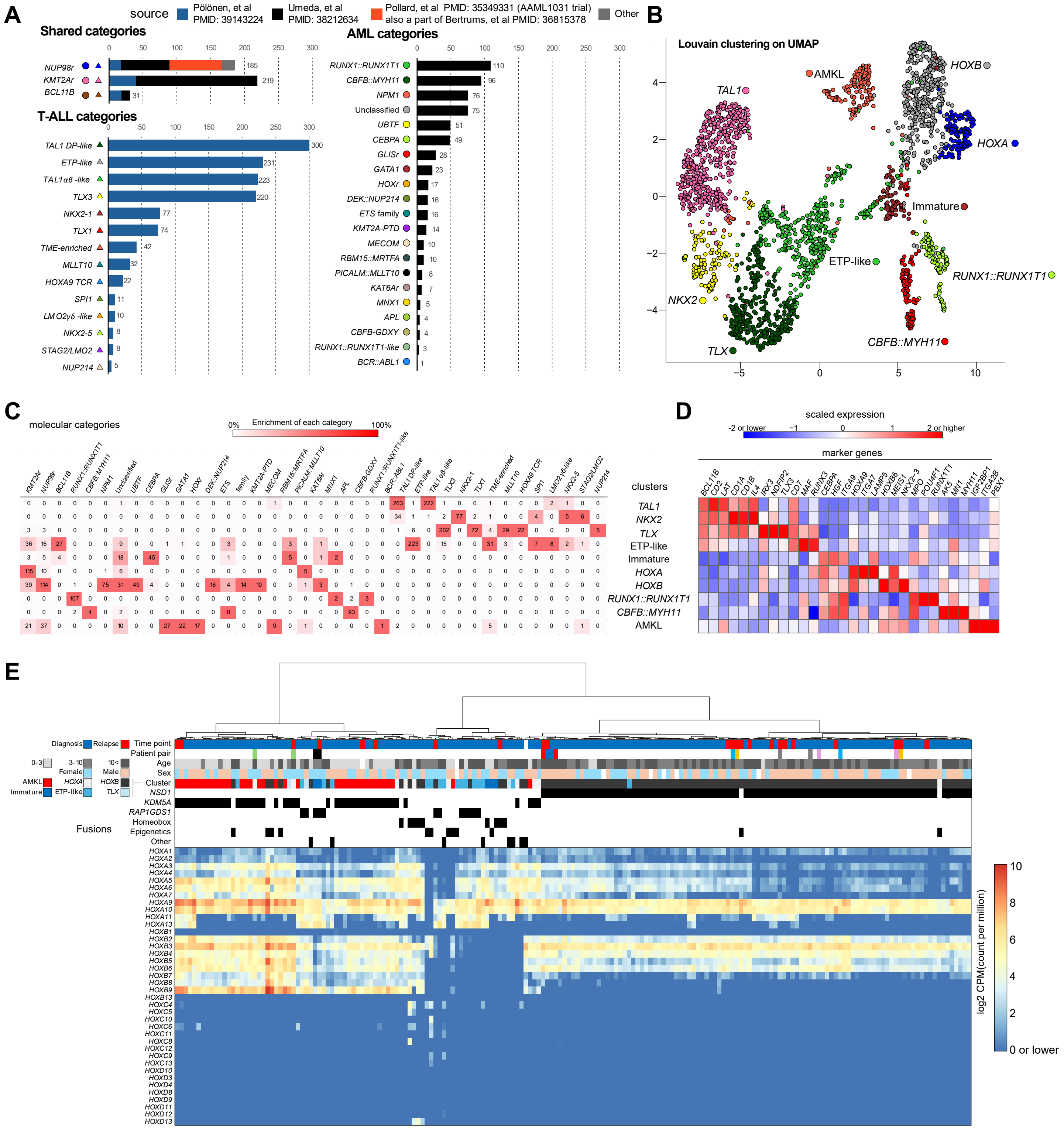


**Figure S2. *Transcriptome analyses with other AML and T-ALL subtypes***

**A**. Numbers of subtypes of T-ALL, AML, or shared subtypes included in the transcriptome analyses (n=2,321). Bars are colored according to source publication listed on the top. **B**. UMAP plot of the transcriptional cohort colored by Louvain clustering. Annotations for clusters are based on major subtypes in the cluster or known marker gene expression. **C**. Enrichment of each subtype in clusters, with colors representing fraction of categories in each cluster. **D**. Heatmap showing marker gene expression in each cluster. Colors indicate scaled expression levels among clusters. **E**. Heatmap showing *HOXA-D* expression in *NUP98*r samples (n=185). Annotations on the top show patient characteristics, and colors of the heatmap show expression levels (log2 CPM: count per million). Samples are clustered according to *HOXA-B* expression levels using Euclid distance and the Ward method.

### Fig.S3

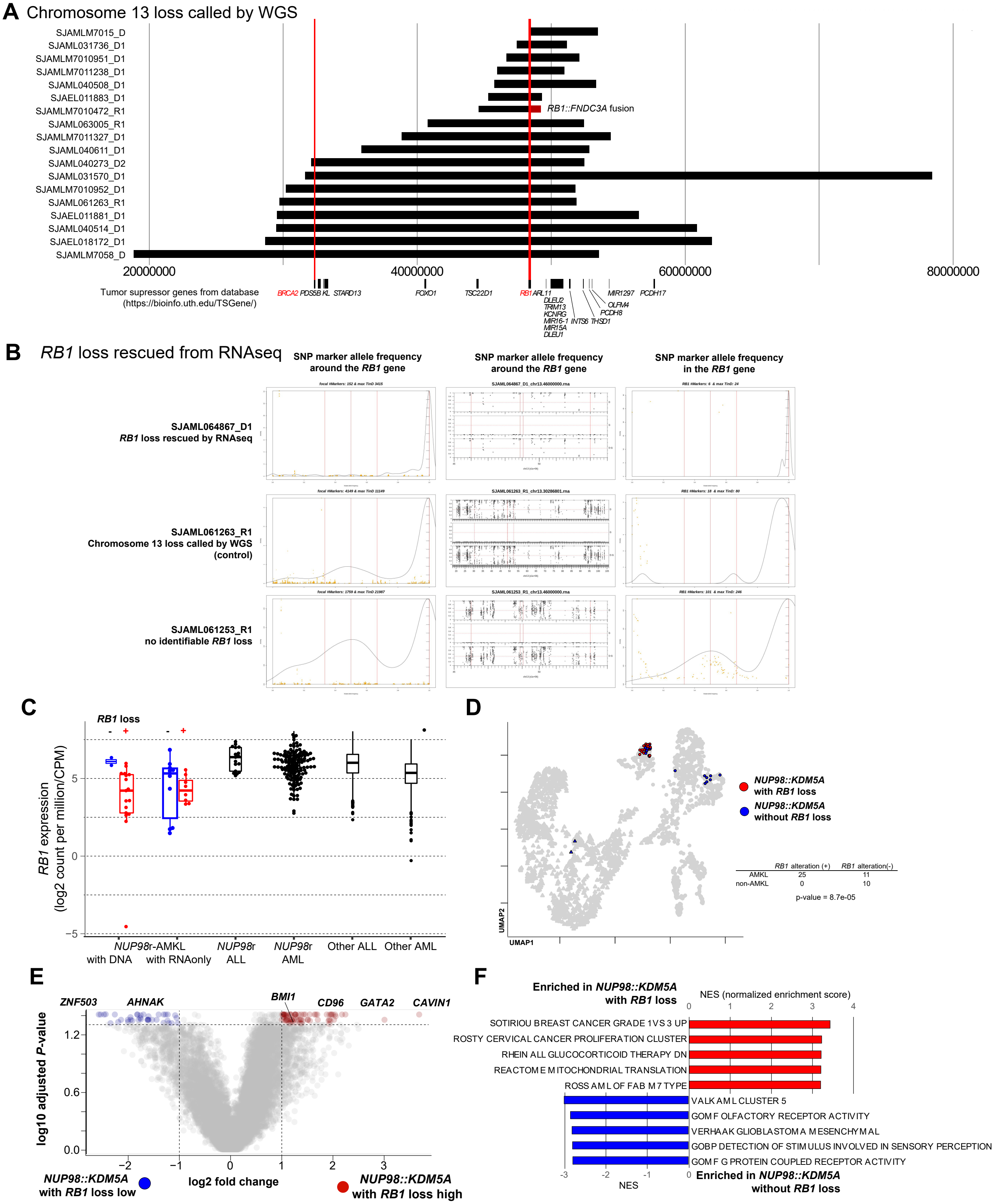
A Chromosome 13 loss called by WGS
SJAMLM7015_D
SJAML031736_D1
SJAMLM7010951_D1
SJAMLM7011238_D1
SJAML040508_D1
SJAEL011883_D1
SJAMLM7010472_R1
SJAML063005_R1
SJAMLM7011327_D1
SJAML040611_D1
SJAML040273_D2
SJAML031570_D1
SJAMLM7010952_D1
SJAML061263_R1
SJAEL011881_D1
SJAML040514_D1
SJAEL018172_D1
SJAMLM7058_D
RB1::FNDC3A fusion
20000000
40000000
60000000
80000000
Tumor supressor genes from database
(https://bioinfo.uth.edu/TSGene/)
BRCA2 PDS5B KL STARD13
FOXO1
TSC22D1
RB1 ARL11
DLEU2
TRIM13
KCNRG
MIR16-1
MIR15A
DLEU1
INTS6
THSD1
PCDH8
OLFM4
MIR1297
PCDH17
B RB1 loss rescued from RNAseq
SNP marker allele frequency around the RB1 gene
SNP marker allele frequency around the RB1 gene
SNP marker allele frequency in the RB1 gene
SJAML064867_D1
RB1 loss rescued by RNAseq
SJAML061263_R1
Chromosome 13 loss called by WGS
(control)
SJAML061253_R1
no identifiable RB1 loss
C
RB1 loss
- +
- +
RB1 expression
(log2 count per million/CPM)
5
0
−5
NUP98r-AMKL
with DNA
with RNAonly
NUP98r
ALL
NUP98r
AML
Other ALL
Other AML
D
NUP98::KDM5A
with RB1 loss
NUP98::KDM5A
without RB1 loss
RB1 alteration (+)
RB1 alteration(-)
AMKL 25 11
non-AMKL 0 10
p-value = 8.7e-05
UMAP2
UMAP1
E
ZNF503
AHNAK
BMI1
CD96
GATA2
CAVIN1
log10 adjusted P-value
1.2
0.6
0.0
−2
−1
0
1
2
3
log2 fold change
NUP98::KDM5A
with RB1 loss low
NUP98::KDM5A
with RB1 loss high
F
Enriched in NUP98::KDM5A
with RB1 loss
NES (normalized enrichment score)
0 1 2 3 4
SOTIRIOU BREAST CANCER GRADE 1 VS 3 UP
ROSTY CERVICAL CANCER PROLIFERATION CLUSTER
RHEIN ALL GLUCOCORTICOID THERAPY DN
REACTOME MITOCHONDRIAL TRANSLATION
ROSS AML OF FAB M7 TYPE
VALK AML CLUSTER 5
GOMF OLFACTORY RECEPTOR ACTIVITY
VERHAAK GLIOBLASTOMA MESENCHYMAL
GOBP DETECTION OF STIMULUS INVOLVED IN SENSORY PERCEPTION
GOMF G PROTEIN COUPLED RECEPTOR ACTIVITY
-3 -2 -1 0
NES
Enriched in NUP98::KDM5A
without RB1 loss

**Figure S3.** ***Characteristics of NUP98::KDM5A leukemia with chromosome 13 loss involving the RB1 locus***

**A**. Chromosomal deletions involving the *RB1* locus in patient samples (n=18). Deleted regions called from whole-genome/exome sequencing (WGS/WES) data are indicated by black bars, and a region inferred from fusion transcripts is indicated by a red bar. **B**. Representative single nucleotide polymorphism (SNP) data used to rescue *RB1* loss in samples with RNAseq data only. SNPs abound the *RB1* genes (**left**), extended area of chromosome 13 (**mid**), and within the *RB1* gene locus (**right**) from a possible case with RNAseq data only (SJAML064868_D1, **top**), a case with *RB1* loss called by WGS (SJAML061263_R1, **mid**), and a case without identifiable *RB1* loss (SJAML061253_R1, **bottom**) are shown as dots. **C**. Expression levels of *RB1* in *NUP98*r leukemia and other leukemia subtypes. Within the *NUP98*r AMKL group, samples are subdivided into those with or without DNA data or *RB1* loss call. Lines of the box plots represent 25% quantile, median, and 75% quantile, the upper whisker represents the higher value of maxima or 1.5 x interquartile range (IQR), and the lower whisker represents the lower value of minima or 1.5 x IQR. Individual data from other ALL or AML are omitted. **D**. UMAP plot of *NUP98::KDM5A* with *RB1* loss (red) or without *RB1* loss (blue). Enrichment of *NUP98::KDM5A* cases with *RB1* in the AMKL cluster was tested using Fisher's exact test (*P*-value = 8.7e-5). **E**. Differentially expressed gene (DEG) analysis between *NUP98::KDM5A* cases within the AMKL cluster with or without *RB1* deletion. Colors indicate DEGs (red: high in cases with *RB1* loss, blue: low in cases without *RB1* loss). **F**. Gene Set Enrichment Analysis (GSEA) between *NUP98::KDM5A* cases within the AMKL cluster with or without *RB1* deletion. Red bars indicate normalized enrichment score (NES) in cases with *RB1* loss, and blue bars indicate NES in cases without *RB1* loss.

### Fig.S4

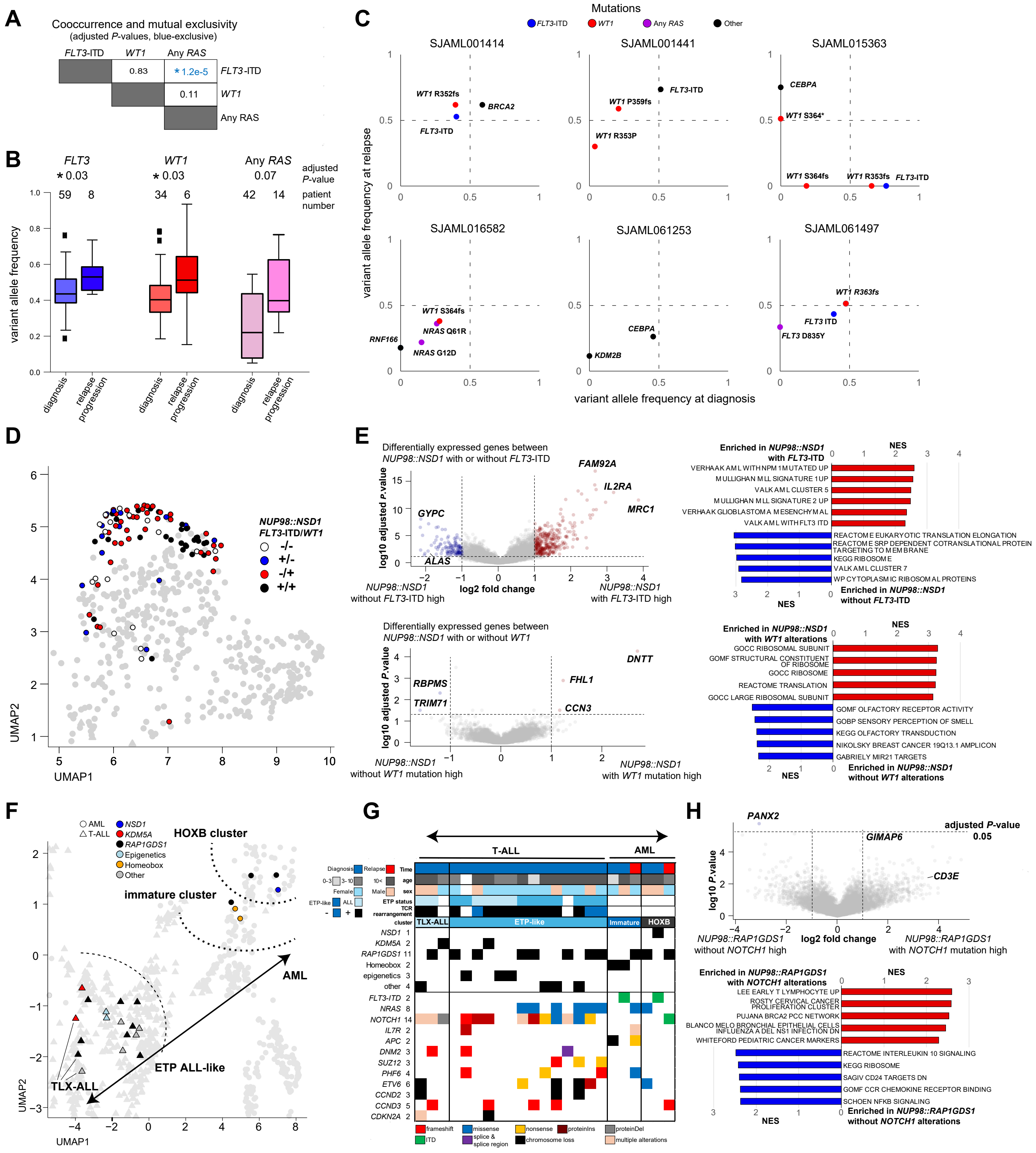
A
Cooccurrence and mutual exclusivity
(adjusted P-values, blue-exclusive)
FLT3-ITD
WT1
Any RAS
0.83
* 1.2e-5
0.11
B
FLT3
WT1
Any RAS
* 0.03
* 0.03
0.07
adjusted P-value
59
8
34
6
42
14
patient number
variant allele frequency
diagnosis
relapse progression
C
Mutations
FLT3-ITD
WT1
Any RAS
Other
SJAML001414
SJAML001441
SJAML015363
SJAML016582
SJAML061253
SJAML061497
WT1 R352fs
BRCA2
FLT3-ITD
WT1 P359fs
WT1 R353P
CEBPA
WT1 S364*
WT1 S364fs
WT1 R353fs
WT1 S364fs
NRAS Q61R
RNF166
NRAS G12D
CEBPA
KDM2B
WT1 R363fs
FLT3 ITD
FLT3 D835Y
variant allele frequency at relapse
variant allele frequency at diagnosis
D
NUP98::NSD1
FLT3-ITD/WT1
-/-
+/-
-/+
+/+
UMAP1
UMAP2
E
Differentially expressed genes between
NUP98::NSD1 with or without FLT3-ITD
FAM92A
IL2RA
MRC1
GYPC
ALAS
log10 adjusted P.value
log2 fold change
NUP98::NSD1 without FLT3-ITD high
NUP98::NSD1 with FLT3-ITD high
Enriched in NUP98::NSD1 with FLT3-ITD
NES
VERHAAK AML WITH NPM1 MUTATED UP
MULLIGHAN MLL SIGNATURE 1 UP
VALK AML CLUSTER 5
MULLIGHAN MLL SIGNATURE 2 UP
VERHAAK GLIOBLASTOMA MESENCHYMAL
VALK AML WITH FLT3 ITD
REACTOME EUKARYOTIC TRANSLATION ELONGATION
REACTOME SRP DEPENDENT COTRANSLATIONAL PROTEIN TARGETING TO MEMBRANE
KEGG RIBOSOME
VALK AML CLUSTER 7
WP CYTOPLASMIC RIBOSOMAL PROTEINS
Enriched in NUP98::NSD1 without FLT3-ITD
Differentially expressed genes between
NUP98::NSD1 with or without WT1
DNTT
FHL1
CCN3
RBPMS
TRIM71
log10 adjusted P.value
NUP98::NSD1 without WT1 mutation high
NUP98::NSD1 with WT1 mutation high
Enriched in NUP98::NSD1 with WT1 alterations
GOCC RIBOSOMAL SUBUNIT
GOMF STRUCTURAL CONSTITUENT OF RIBOSOME
GOCC RIBOSOME
REACTOME TRANSLATION
GOCC LARGE RIBOSOMAL SUBUNIT
GOMF OLFACTORY RECEPTOR ACTIVITY
GOBP SENSORY PERCEPTION OF SMELL
KEGG OLFACTORY TRANSDUCTION
NIKOLSKY BREAST CANCER 19Q13.1 AMPLICON
GABRIELY MIR21 TARGETS
Enriched in NUP98::NSD1 without WT1 alterations
F
AML
T-ALL
NSD1
KDM5A
RAP1GDS1
Epigenetics
Homeobox
Other
HOXB cluster
immature cluster
AML
ETP ALL-like
TLX-ALL
UMAP1
UMAP2
G
T-ALL
AML
Diagnosis
Relapse
Time
age
sex
Female
Male
ETP status
ETP-like
ALL
TCR rearrangement
cluster
TLX-ALL
ETP-like
Immature
HOXB
NSD1 1
KDM5A 2
RAP1GDS1 11
Homeobox 2
epigenetics 3
other 4
FLT3-ITD 2
NRAS 8
NOTCH1 14
IL7R 2
APC 2
DNM2 3
SUZ12 3
PHF6 4
ETV6 6
CCND2 3
CCND3 5
CDKN2A 2
frameshift
missense
nonsense
proteinIns
proteinDel
ITD
splice & splice region
chromosome loss
multiple alterations
H
PANX2
GIMAP6
CD3E
adjusted P-value
0.05
log10 P.value
log2 fold change
NUP98::RAP1GDS1 without NOTCH1 high
NUP98::RAP1GDS1 with NOTCH1 mutation high
Enriched in NUP98::RAP1GDS1 with NOTCH1 alterations
NES
LEE EARLY T LYMPHOCYTE UP
ROSTY CERVICAL CANCER PROLIFERATION CLUSTER
PUJANA BRCA2 PCC NETWORK
BLANCO MELO BRONCHIAL EPITHELIAL CELLS INFLUENZA A DEL NS1 INFECTION DN
WHITEFORD PEDIATRIC CANCER MARKERS
REACTOME INTERLEUKIN 10 SIGNALING
KEGG RIBOSOME
SAGIV CD24 TARGETS DN
GOMF CCR CHEMOKINE RECEPTOR BINDING
SCHOEN NFKB SIGNALING
Enriched in NUP98::RAP1GDS1 without NOTCH1 alterations

**Figure S4. *Characteristics of NUP98::NSD1 or NUP98::RAP1GDS1 leukemia with cooperating alterations***

**A**. Co-occurrence of *FLT3*-ITD, *WT1*, or any RAS-related mutations among *NUP98::NSD1* cases, with blue adjusted *P*-value indicating exclusivity. **B**. Comparison of variant allele frequencies (VAF) of *FLT3*-ITD, *WT1*, or any RAS-related mutations at diagnosis and relapse/progression. **C**. Comparison of VAFs between two time points of *NUP98::NSD1* cases. Colors of dots indicate types of mutations. **D**. Distributions of *NUP98::NSD1* cases on the UMAP plot. The colors of dots indicate the status of *FLT3*-ITD and *WT1* mutations. **E**. DEG analyses (**left**) and GSEA (**right**) between *NUP98::NSD1* cases with or without *FLT3*-ITD (**top**) or *WT1* mutations (**bottom**). Red dots and bars indicate DEGs or GO terms enriched in cases with *FLT3*-ITD or *WT1* mutations. **F**. Distributions of immature ~ T-ALL *NUP98*r leukemia cases on the UMAP plot. The colors of dots indicate fusion groups, and the shapes indicate disease types. **G**. Mutational heatmap of immature to T-ALL *NUP98*r cases. Colors of annotations (**top**) indicate patient characteristics, and colors of heatmap indicate mutation types (**bottom**). **H**. DEG analysis of *NUP98::RAP1GDS1* cases with or without *NOTCH1* mutations (top) and GSEA (bottom). Only one gene (*PANX2*) low in *NOTCH1*+ cases was identified. Statistical tests were performed by two-sided Fisher's exact test (**A**), the Wilcoxon rank sum test (**B,** two-sided) and limma (**E**,**H**) followed by the Benjamini-Hochberg adjustment.

### Fig.S5

**A**

nFeature_RNA > 200 & percent.mt < 20

| sample | fusion | disease | before fitering | after filtering | pass | |
|---|---|---|---|---|---|---|
| SJAML015363_D5 | *NUP98::NSD1* | AML | 3547 | 2511 | 70.8% | |
| SJAML016582_D1 | *NUP98::NSD1* | AML | 4335 | 2478 | 57.2% | |
| SJAML061252_R1 | *NUP98::NSD1* | AML | 4576 | 3086 | 67.4% | |
| SJAML001441_D3 | *NUP98::NSD1* | AML | 3181 | 2130 | 67.0% | |
| SJAML001441_R4 | *NUP98::NSD1* | AML | 3175 | 2314 | 72.9% | |
| PAXLWH_D | *NUP98::NSD1* | AML | 1413 | 1413 | 100.0% | prefiltered |
| PAXLWH_R | *NUP98::NSD1* | AML | 8058 | 8058 | 100.0% | |
| SJAML031254_D1 | *NUP98::RAP1GDS1* | AML (immature) | 206 | 188 | 91.3% | |
| SJTALL005_D | *NUP98::RAP1GDS1* | ALL | 76 | 64 | 84.2% | |
| SJALL040121_D1 | *NUP98::KDM5A* | ALL | 147 | 112 | 76.2% | |
| SJAML061263_R1 | *NUP98::KDM5A* | AMKL | 889 | 830 | 93.4% | |

**B**

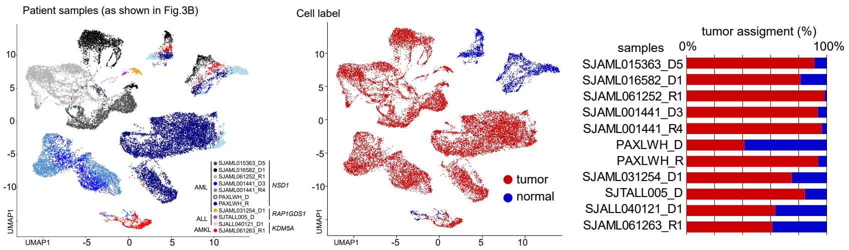


**C**

**D**

**E**

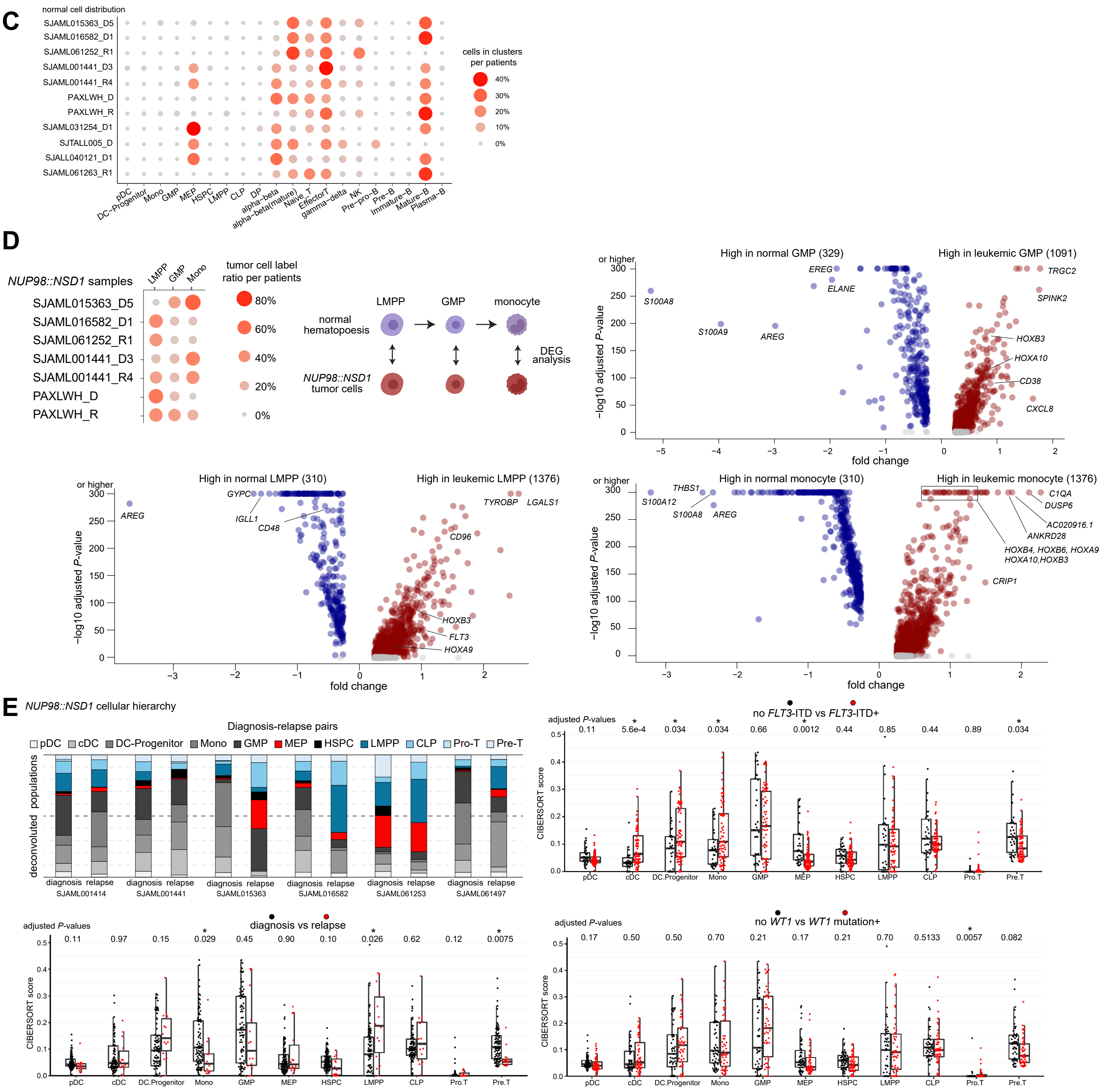

**Figure S5.** ***Single cell and CIBERSORT profiling of NUP98::NSD1 cases***

**A**. Table showing single cell RNAseq (scRNAseq) data passed through 10X pipeline and additional filtering using Seurat (nFeature_RNA > 200 & percent.mt < 20). Public data from COG (Children's Oncology Group) has been prefiltered. **B.** UMAP plots of scRNAseq data colored by Louvain clustering (**left**, as shown in Fig.3B), or by tumor labels based on the clustering (**mid**), and tumor fractions in each patient sample (**right**). **C**. Enrichment of normal cells with each cell label inferred by Seurat, indicated by colors and sizes. **D**. The enrichment of cells from *NUP98::NSD1* samples labeled as LMPP, GMP, and monocyte-like (extracted from Fig.3F), and schema showing DEG design between normal hematopoiesis and *NUP98::NSD1* cases (**top-left**), DEG analyses between normal and leukemic LMPP populations (**left-bottom**) and normal and leukemic GMP populations (**top-right**), and normal and leukemic monocyte population (**bottom-right**). Red dots indicate DEGs high in leukemic populations, and blue dots indicate DEGs high in normal populations. **E**. Comparison of cellular hierarchies of *NUP98::NSD1* cases between different time points (**top-left**), samples at diagnosis or relapse (**bottom-left**), with or without *FLT3*-ITD (**top-right**), and with or without *WT1* mutations (**bottom-right**). In **D**, DEGs were identified using FindMarker function in Seurat package with default settings, which calculate adjusted *P*-values with limma implementation of the Wilcoxon rank-sum test followed by Bonferroni correction. For those with adjusted *P*-values lower than 1.0e-300, exact values were not calculated and plotted as *P*=1.0e-300. Statistical tests in **E** were performed by the Wilcoxon rank sum test followed by the Benjamini-Hochberg adjustment.

Fig.S6

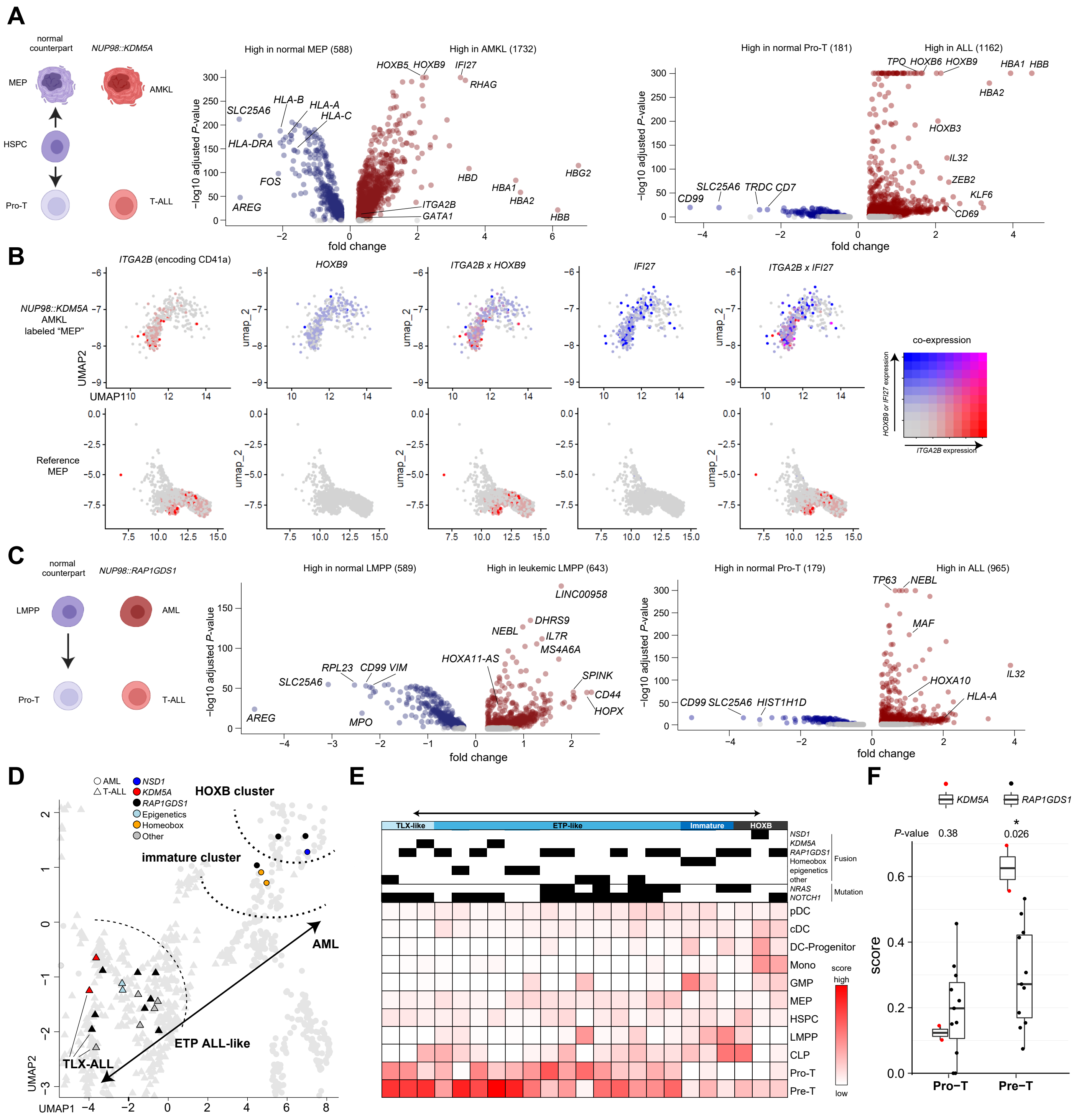


**Figure S6. *Single cell and CIBERSORT profiling of NUP98::KDM5A and ::RAP1GDS1 cases***

**A**. Schema showing cellular hierarchy in normal hematopoiesis and *NUP98::KDM5A* cases (**left**), DEG analyses between normal and leukemic MEP (**mid**) and normal and leukemic Pro-T populations (**right**). Red dots indicate DEGs high in leukemic populations, and blue dots indicate DEGs high in normal populations. **B**. Co-expression profiles of *ITGA2B* (encoding CD41a) and *HOXB9* **(mid)** and *IFI27* (**right**) among normal MEP cells and AMKL cells projected onto the reference UMAP. The colors of the dot indicate gene expression (red: *ITGA2B*, blue: *HOXB9* or *IFI27,* purple: co-expression, gray: no expression). **C**. Schema showing cellular hierarchy in normal hematopoiesis and *NUP98::RAP1GSD1* cases (**left**), DEG analyses between normal and leukemic LMPP (**mid**) and normal and leukemic Pro-T populations (**right**) as shown in **A**. **D**. Distributions of immature ~ T-ALL *NUP98*r leukemia cases on the UMAP the plot as shown in Fig.S4F. The colors of dots indicate fusion groups, and the shapes indicate disease types. **E**. Cellular hierarchies of immature to T-ALL *NUP98*r cases inferred by CIBERSORT, with colors indicating signature intensity. **F**. Comparison of Pro-T and Pre-T scores between *NUP98::KDM5A* and *NUP98::RAP1GDS1* cases in these immature ~ T-ALL clusters. In **A** and **C**, DEGs were identified using FindMarker function in Seurat package with default settings, which calculate adjusted *P*-values with limma implementation of the Wilcoxon rank-sum test followed by Bonferroni correction. For those with adjusted *P*-values lower than 1.0e-300, exact values were not calculated and plotted as *P*=1.0e-300. In **F**, statistical tests were performed by the Wilcoxon rank sum test followed by the Benjamini-Hochberg adjustment.

### Fig.S7

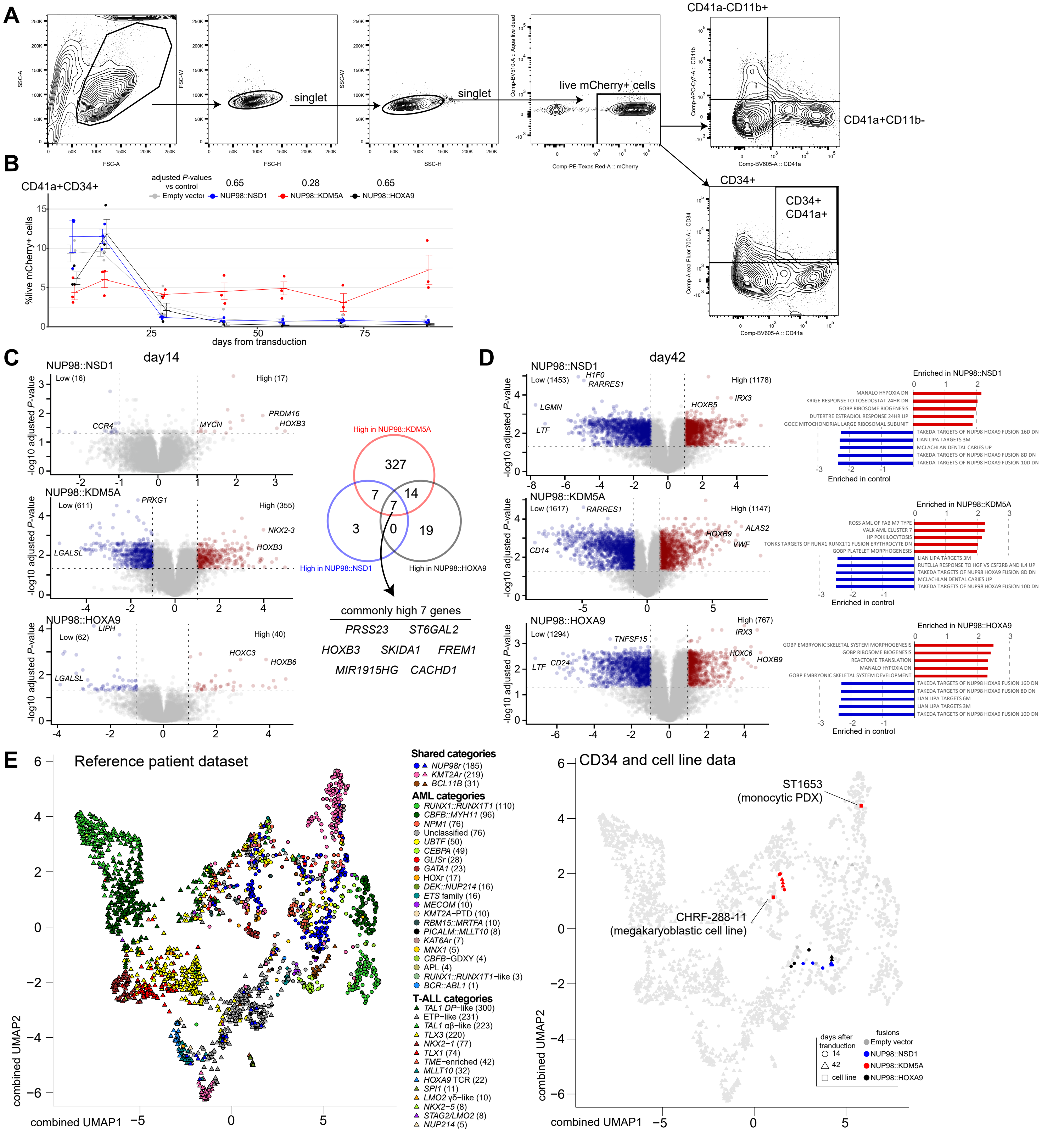

**Figure S7. *Cord blood CD34+ cell (cbCD34) models of NUP98r leukemia***

**A**. Gating strategy for flow cytometric analyses of cbCD34 models in liquid culture. **B**. Flow cytometric analysis of CD41a+CD34+ population in cbCD34 models in liquid culture. **C**. Comparison of DEGs in each cbCD34 model compared with empty vector controls at day 14 (**top**: NUP98::NSD1, **mid**: NUP98::KDM5A, **bottom**: NUP98::HOXA9). Colors indicate high (red) or low (blue) in each CD34 model. Venn diagram shows overlapping genes high in each model (**right**). **D**. DEGs (**left**) as shown in **C** and GSEA (**right**) of cbCD34 model compared with empty vector controls at day 42 (**right**). **E**. UMAP plots of a combined cohort of patient samples in Fig.1D (n=2,321) and cbCD34 models in Fig.3E (n=26). Colors of dots indicate molecular subtypes of reference data (**left**) and NUP98 FOs (**right**), and shapes indicate days from transduction. Data was obtained from three biological replicates (different lots of cord blood). Statistical tests were performed by linear mixed effect model followed by comparison with empty vector control (**B**) or limma (**C**, **D**) and correction with the Benjamini-Hochberg adjustment. Error bars indicate mean ± s.e.m.

### Fig.S8

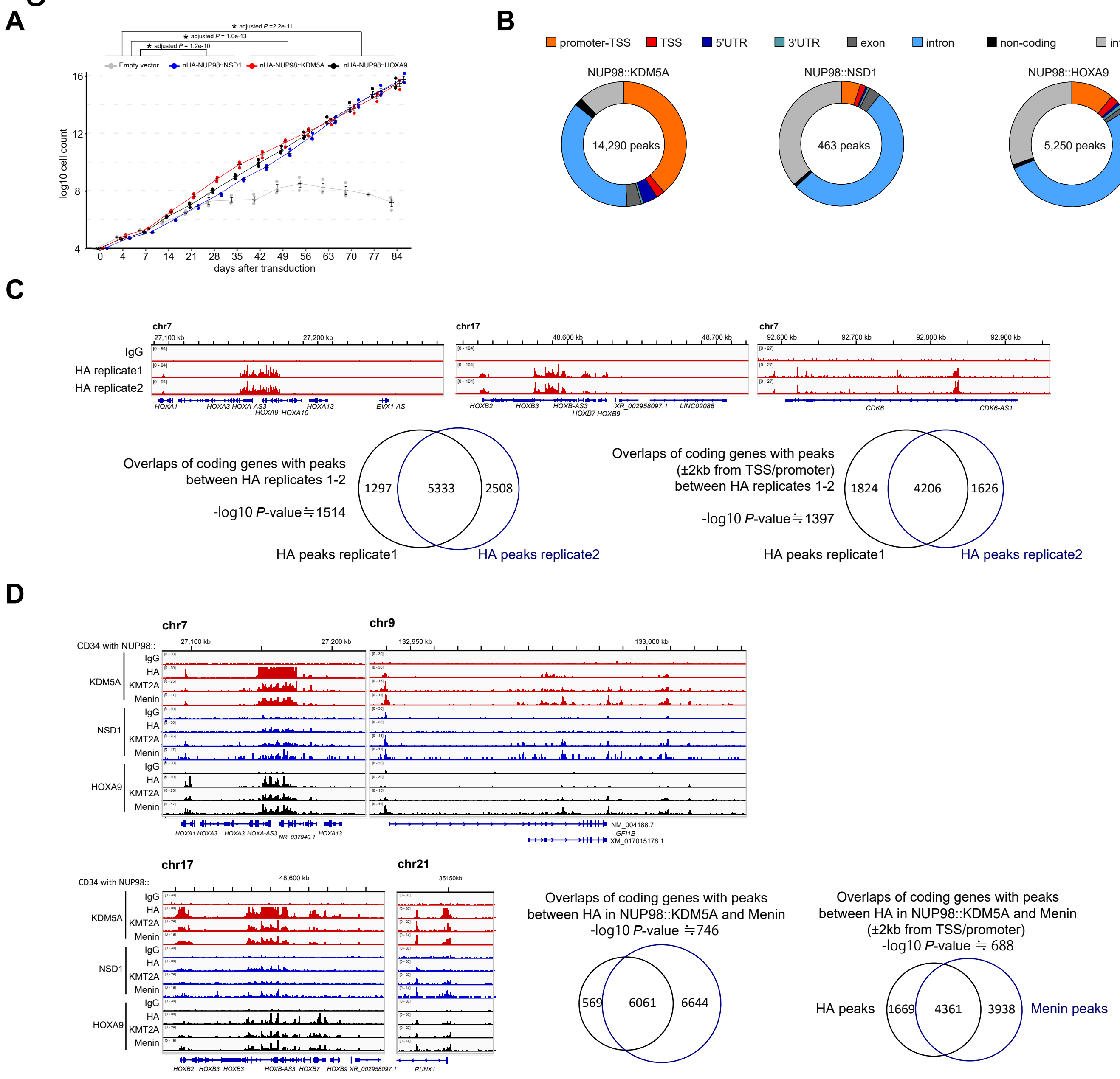


**Figure S8.** ***Epigenetic profiling of NUP98r cbCD34 models***

**A**. Cell growth assays of cbCD34 models with N-terminus HA-tagged NUP98 fusions in liquid culture. **B**. Annotation of peaks from CUT&RUN using HA antibodies in HA-tagged NUP98-FO cbCD34 models (**left**-NUP98::KDM5A, **mid**-NUP98::NSD1, **right**-NUP98::HOXA9). Colors indicate peak annotations. **C**. IGV tracks at *HOXA* cluster (**top-left**), the *HOXB* cluster (**top-mid**), and *CDK6* (**top-right**) comparing two biological replicates of HA-tagged NUP98::KDM5A cbCD34 model, and Venn diagrams comparing overlaps of target protein-coding genes with all peaks (**bottom-left**) and with peaks within 2kb from TSS/promoter (**bottom-right**). **D**. IGV tracks at the *HOXA* cluster (**top-left**), *GFI1* (**top-right**), the *HOXB* cluster (**bottom-left**), and the *RUNX1* promoter (**bottom-mid**) comparing HA, KMT2A, and menin signals from HA-tagged cbCD34 models, and Venn diagrams (**bottom-right**) comparing overlaps of protein-coding genes with HA and menin peaks (all peaks and peaks within 2kb from TSS/promoter). In **A**, statistical tests were performed by linear mixed effect model followed by the Benjamini-Hochberg adjustment, asterisks indicating adjusted *P*-values *<0.05. Error bars indicate mean ± s.e.m. Abbreviations. Statistical significances of target gene overlaps were assessed with a hypergeometric test (one-sided). For those with *P*-values lower than 1.0e-320, exact values were not calculated, and proximal values are shown. Abbreviations. TSS: transcription start site, UTR: untranslated region

### Fig.S9

A

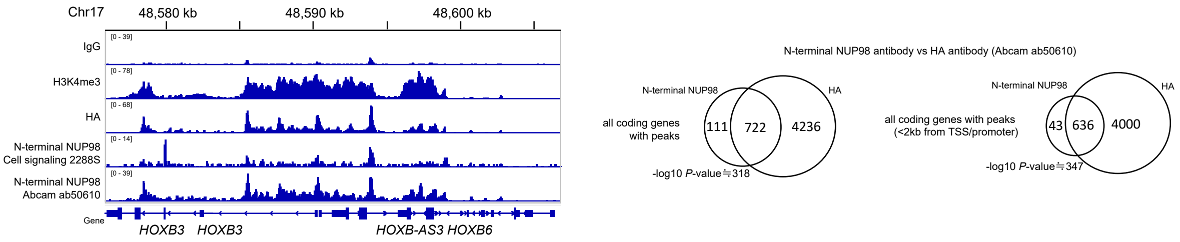


B

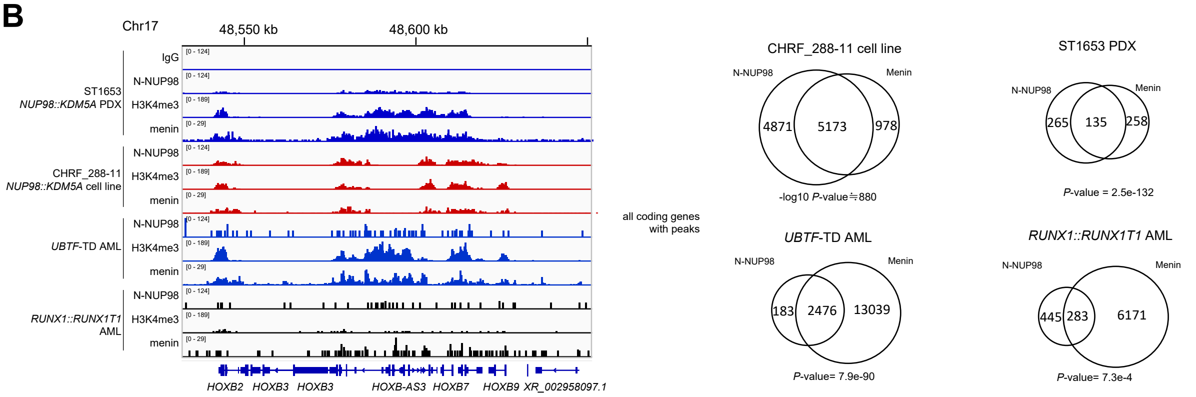


C

| patient samples | fusions | cluster | antibody: H3K4me3 | H3K27Ac | H3K27me3 | N-terminal NUP98 |
|---|---|---|---|---|---|---|
| SJAML031736_D4 | *KDM5A* | AMKL | | | | |
| SJAML061263_R4 | *KDM5A* | AMKL | | | | |
| SJAML031570_D2 | *KDM5A* | AMKL | | | | |
| SJAML016570_D4 | *KDM5A* | AML | | | | |
| SJAML016570_R2 | *KDM5A* | AML | | | | |
| SJALL040121_D3 | *KDM5A* | T-ALL | | | | excluded |
| SJAML001441_R7 | *NSD1* | AML | | | | excluded |
| SJAML015363_D6 | *NSD1* | AML | excluded | excluded | excluded | excluded |
| SJAML001441_D4 | *NSD1* | AML | excluded | | | excluded |
| SJAML031434_D4 | *NSD1* | AML | | | | |
| SJAML015347_D3 | *NSD1* | AML | | | | |
| SJAML001409_D3 | *RAP1GDS1* | immature | excluded | excluded | excluded | excluded |
| SJAML001409_R2 | *RAP1GDS1* | immature | | | | |
| SJAML031254_D6 | *RAP1GDS1* | immature | | | | |

D

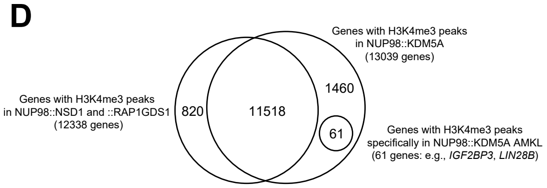


E

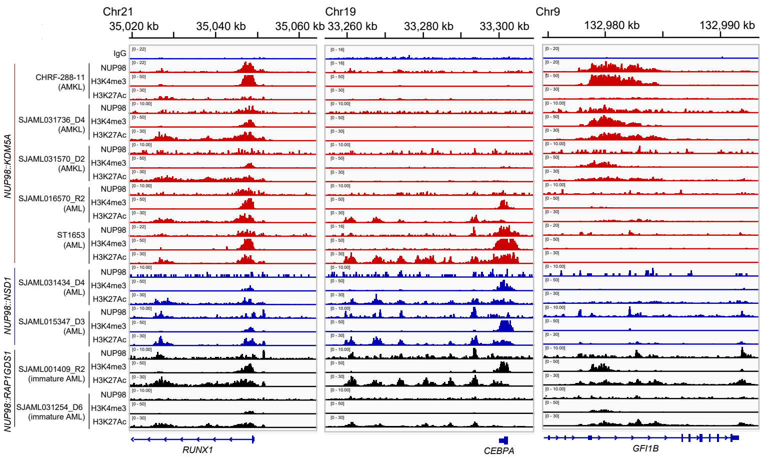


**Figure S9. *Epigenetic profiling of NUP98r primary and cell line samples***

**A**. IGV tracks at the *HOXB* cluster (**left**) comparing CUT&RUN signals using HA and two N-NUP98 antibodies in HA-tagged NUP98::KDM5A cbCD34 model, and Venn diagrams comparing overlaps target protein-coding genes with all peaks (**mid**) and with peaks within 2kb from TSS/promoter (**right**) from HA and N-NUP98 (ab50620) antibodies. **B**. IGV tracks at the *HOXB* cluster (**left**) comparing N-NUP98, H3K4me3, and menin signals from ST1653 (*NUP98::KDM5A* PDX, myelomonocytic), CHRF-288-11 (*NUP98::KDM5A* cell line, megakaryocytic), and primary AML of other subtypes (*UBTF*-tandem duplication: TD , *RUNX1::RUNX1T1*), and Venn diagrams (**right**) comparing overlaps of protein-coding genes with N-NUP98 and menin peaks in each sample. **C**. Lists of primary *NUP98*r leukemia samples profiled with CUT&RUN. Data with high background or low signals from assay conditions was excluded from the following analyses. **D**. Overlap of genes with H3K4me3 peaks among non-NUP98::KDM5A and NUP98::KDM5A. **E**. IGV tracks of signals from CUT&RUN using N-terminal NUP98, H3K4me3, and H3K27ac antibodies in primary leukemia samples and NUP98::KDM5A cell lines on *RUNX1* (**left**: shared target), *CEBPA* (**mid**: non-AMKL target), and *GFI1B* (**right**: AMKL target). Statistical significances of target gene overlaps were assessed with hypergeometric test (one-sided). For those with *P*-values lower than 1.0e-320, exact values were not calculated, and proximal values are shown.

### Fig.S10

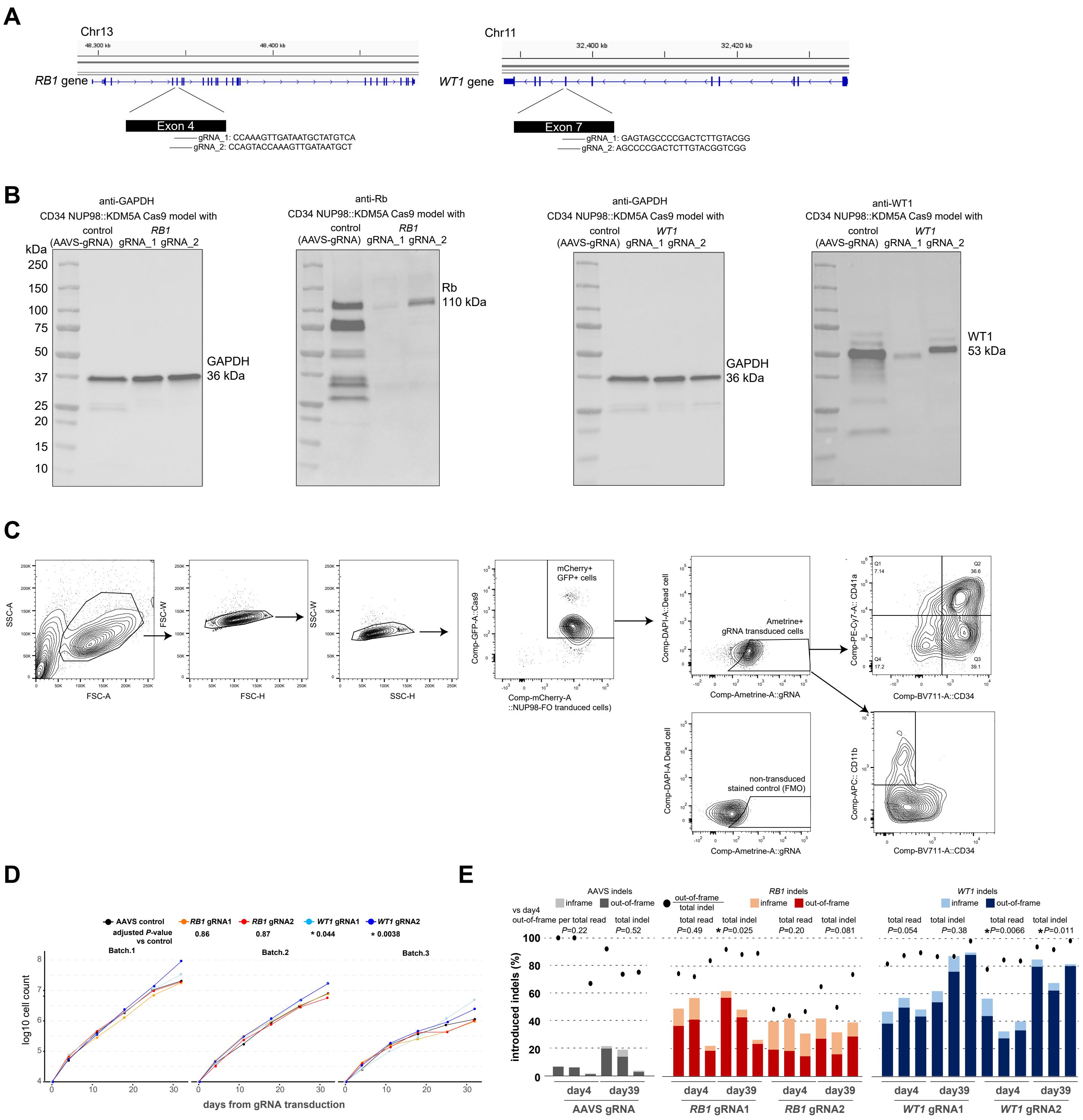


**Figure S10.** ***Modeling of cooperating alterations in cbCD34 models***

**A**. Locations and sequences of gRNA designed to mimic functional *RB1* loss (**left**: truncation at exon 4) and recurrent *WT1* mutations (**right**: frameshift at exon 7). **B.** Western blotting for RB1 (**left**) and WT1 (**right**) proteins in cord blood CD34 Cas9 models with gRNA. **C**. Gating strategies for gRNA transduced cells with mAmetrine expression. **D**. Cell growth assays of cbCD34/Cas9 *NUP98::NSD1* line with gRNAs targeting the AAVS, *RB1*, or *WT1* loci. Due to decline in cell growth over time, each replicate is shown separately. **E**. Induction rates of indel (insertions and deletions) at day 4 and 32 in NUP98::NSD1/Cas9 line in each condition. Bars represent fractions of indel rates in all target sequence reads, and dots represent out-of-frame indel ratio among total indels. Data was obtained in technical triplicates from an established NUP98::NSD1/Cas9 line and independent experiments. Statistical tests were performed by generalized linear mixed effect model with Gaussian distributions including batches in a model followed by the Benjamini-Hochberg adjustment (**C**) and Student's t-test by comparing day 4 and day 32 (**D**). Asterisks indicating *P*-values or adjusted *P*-values <0.05.

### Fig.S11

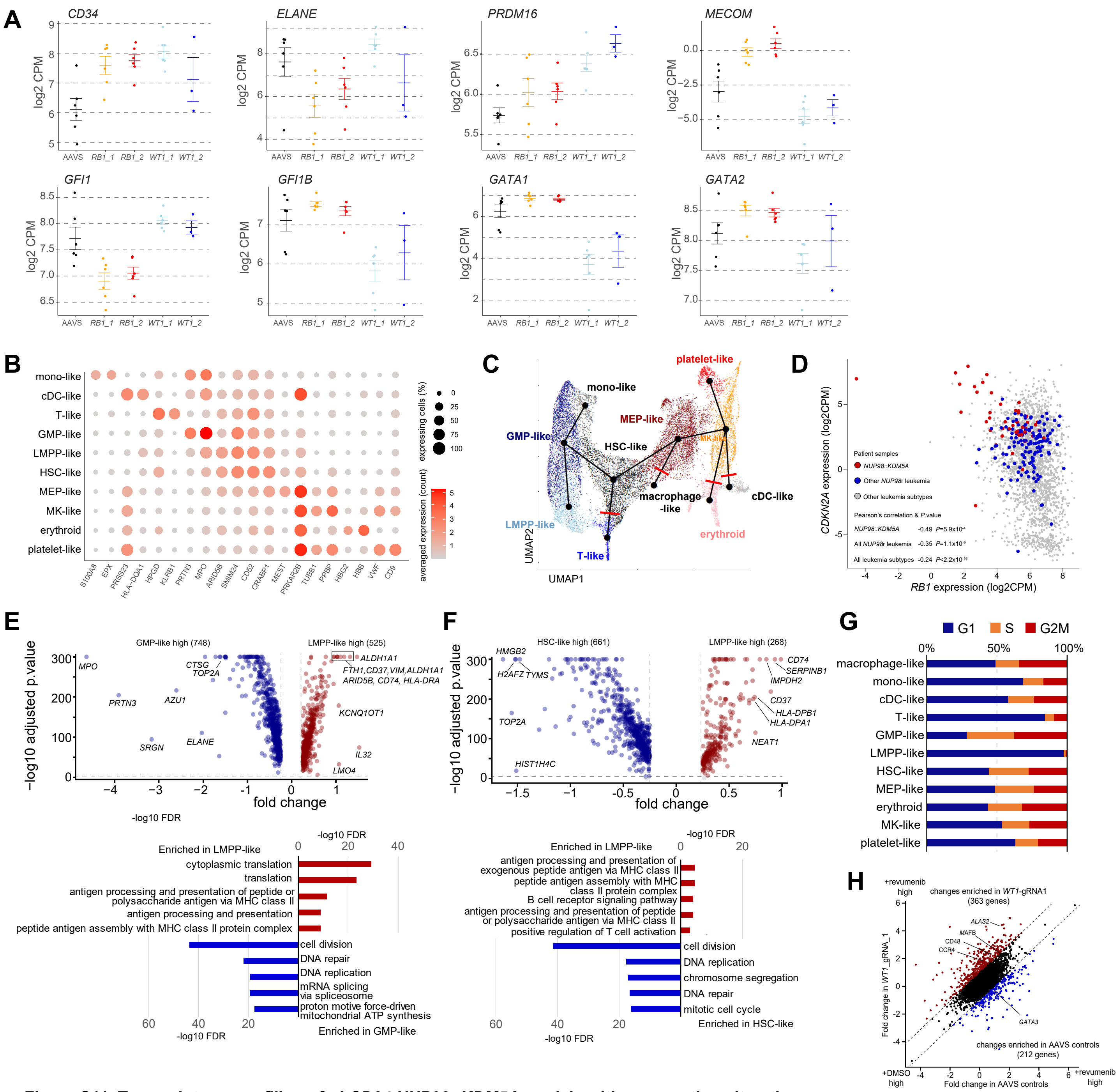

**Figure S11. Transcriptome profiling of *cbCD34 NUP98::KDM5A models with cooperating alterations***

**A**. Expression levels of selected hematopoietic differentiation genes in cbCD34 models with gRNA. Statistical tests were performed using the entire gene set as shown in Fig. 6F-G. **B.** Marker gene expression in each cluster in scRNAseq from gRNA-transduced cbCD34 NUP98::KDM5A models, indicated by colors (averaged expression) and size (ratio of expressing cells: count>0). **C.** Differentiation branches used for pseudotime analyses. Excluded branches are indicated by red bars. **D.** Comparison of expression levels of *RB1* and *CDKN2A* in the bulk RNA-sequence cohort. The colors of dots indicate sample types (red-NUP98::KDM5A, blue-other *NUP98*r leukemia, gray-other leukemia subtypes). Pearson correlations and statistical values (*P*-value) within each group are shown at the bottom. **E.** DEG analysis between the LMPP-like and GMP-like clusters in the *WT1*-gRNA condition (**top**) and GO term analysis (**bottom**) of genes high in the LMPP-like cluster (red) and the GMP-like cluster (blue). **F.** DEG analysis between the LMPP-like and HSC-like clusters in the *WT1*-gRNA condition (**top**) and GO term analysis (**bottom**) of genes high in the LMPP-like cluster (red) and the HSC-like cluster (blue). **G.** Cell cycle status of cells in each cluster inferred by the CellCycleScoring function from the Seurat package. **H.** comparison of expression changes between AAVS and *WT1*-gRNA1 conditions (**left**) and GO term analysis of changes enriched (difference of fold changes >1) in *WT1*-gRNA1 conditions (**right**). In **D-F**, statistical tests were performed by limma followed by the Benjamini-Hochberg adjustment (**E, F**) or Pearson correlation coefficient using the cor.test function.
